# Community interventions for people with complex emotional needs that meet the criteria for ‘personality disorder’ diagnoses: a systematic review of economic evaluations

**DOI:** 10.1101/2020.11.03.20225078

**Authors:** Joe Botham, Amy Clark, Thomas Steare, Ruth Stuart, Sian Oram, Brynmor Lloyd-Evans, Tamar Jeynes, Eva Broeckelmann, Mike Crawford, Sonia Johnson, Alan Simpson, Paul McCrone

**Affiliations:** NIHR Mental Health Policy Research Unit, Health Service and Population Research Department, Institute of Psychiatry, Psychology & Neuroscience, King’s College London; NIHR Mental Health Policy Research Unit, Division of Psychiatry, University College London; Institute for Lifecourse Development, University of Greenwich; NIHR Mental Health Policy Research Unit Complex Emotional Needs Lived Experience Working Group, Health Service and Population Research Department, Institute of Psychiatry, Psychology & Neuroscience, King’s College London; Centre of Psychiatry, Imperial College London

**Author notes:** Address for correspondence: Paul McCrone, Institute for Lifecourse Development, University of Greenwich, Old Royal Naval College, Park Row, Greenwich SE10 9LS. Registered to PROSPERO (ID: CRD42020134068).

## Abstract

**Background:** Diagnoses of “personality disorder” are prevalent among people using community secondary mental health services. Whilst the effectiveness of a range of community-based treatments have been considered, as the NHS budget is finite, it is also important to consider the cost-effectiveness of those interventions.

**Aims:** To assess the cost-effectiveness of primary or secondary care community-based interventions for people with complex emotional needs that meet criteria for a diagnosis of “personality disorder” to inform healthcare policy making.

**Method:** Systematic review (PRESPORO #: CRD42020134068) of five databases, supplemented by reference list screening and citation tracking of included papers. We included economic evaluations of interventions for adults with complex emotional needs associated with a diagnosis of ‘personality disorder’ in community mental health settings published between before 18 September 2019. Study quality was assessed using the CHEERS statement. Narrative synthesis was used to summarise study findings.

**Results:** Eighteen studies were included. The studies mainly evaluated psychotherapeutic interventions. Studies were also identified which evaluated altering the setting in which care was delivered and joint crisis plans. No strong economic evidence to support a single intervention or model of community-based care was identified.

**Conclusion:** There is no robust economic evidence to support a single intervention or model of community-based care for people with complex emotional needs. The review identified the strongest evidence for Dialectical Behavioural Therapy with all three identified studies indicating the intervention is likely to be cost-effective in community settings compared to treatment as usual. Further research is needed to provide robust evidence on the cost-effectiveness of community-based interventions upon which decision makers can confidently base guidelines or allocate resources.

## Introduction

Globally, it is estimated that approximately eight percent of the population experience complex emotional needs that meet the diagnostic criteria for ‘personality disorder’ which is described broadly as an enduring and pervasive pattern of emotional and cognitive difficulties which affect the way in which a person relates to others or understand themselves(1). The diagnosis is often associated with high rates of co-morbidity (2) high levels of service use (3) and high treatment costs (4).

In European and North American community secondary mental healthcare settings, the prevalence of ‘personality disorder’ diagnoses are estimated to be above 40% (5). A range of psychological therapies show some evidence of effectiveness; dialectical behavioural therapy (DBT) and Mentalisation-Based Treatment (MBT) have the most well-established evidence base (6,7). While establishing clinical effectiveness of psychological therapies and models of care is crucial, it is also important for decision makers to consider their value for money. Health and social care resources are limited and, as such, there are competing demands for scarce resources. It is therefore a growing requirement that assessments of new treatments and therapies include an economic evaluation (8). Through robust economic evaluation decision makers can consider the opportunity cost of funding one intervention over another, as for every potential gain from a funded intervention, given a limited funding pool, there are potential losses from the next best alternative option that is forgone (9).

Economic evaluation seeks to compare the costs of an intervention against its outcomes. The main approaches are cost-effectiveness analysis (CEA), cost-benefit analysis (CBA), cost consequences analysis (CCA), or cost-utility analysis (CUA) (10). CEAs compare the incremental cost of an intervention to incremental changes in outcomes using a condition specific measure. For complex emotional needs these may include an assessment of functioning or distress. CBA measures outcomes using monetary units and are uncommon in healthcare evaluations partly due to the challenge of valuing health effects in monetary terms. CCA presents costs against a number of outcome measures to support multi-criterion decision making. Finally, cost-utility analysis (CUA) is a type of CEA and compares the incremental cost of an intervention against changes in a generic measure of health, usually quality-adjusted life years (QALYs) where one QALY is equivalent to one year of life in perfect health. Results can be compared against a willingness to pay (WTP) threshold which indicates how much a society will pay for a QALY. The National Institute for Health and Care Excellence use a threshold of up to £30,000 in England (11).

We are aware of two systematic reviews of economic evaluations of interventions for ‘personality disorder’(3,4). The scope of these reviews was limited to interventions for people with diagnoses of ‘borderline personality disorder’ only and the most recent papers included were published in 2014. The aim of this paper is to assess the cost-effectiveness of community-based interventions for people with complex emotional needs that meet criteria for diagnoses of ‘personality disorder’ compared to usual care (as defined by each study) or other active interventions. To do this we systematically review and assess the quality of relevant economic evaluations.

## Methods

This systematic literature review was carried out in accordance with “The Preferred Reporting items for Systematic Reviews and Meta-Analyses” (PRISMA) checklist(12). A protocol for the search strategy and methods has been registered with PROSPERO (CRD42020134068). We use ‘complex emotional needs’ as our preferred terminology rather than ‘personality disorder’. This choice seeks to recognise that although some people find the diagnostic term helpful, many find it to be invalidating and stigmatising. However, we do use the term ‘personality disorder’ when referring to original papers and search strategy inclusion criteria (diagnostic inclusion criterion for the service model described) as appropriate.

### Eligibility criteria

Inclusion criteria (detailed in Table 1) required that studies (i) included an economic evaluation where both costs and outcomes are reported and where a formal or informal linkage between costs and outcomes is made; (ii) included adults attending a general or specialist community mental health service with complex emotional needs that meet the diagnostic criteria for ‘personality disorder’ (other than ‘antisocial personality disorder’) (iii) evaluated community-based services, treatments and interventions; (iv) compared cost-effectiveness to usual care or another active intervention.

**Table 1.**
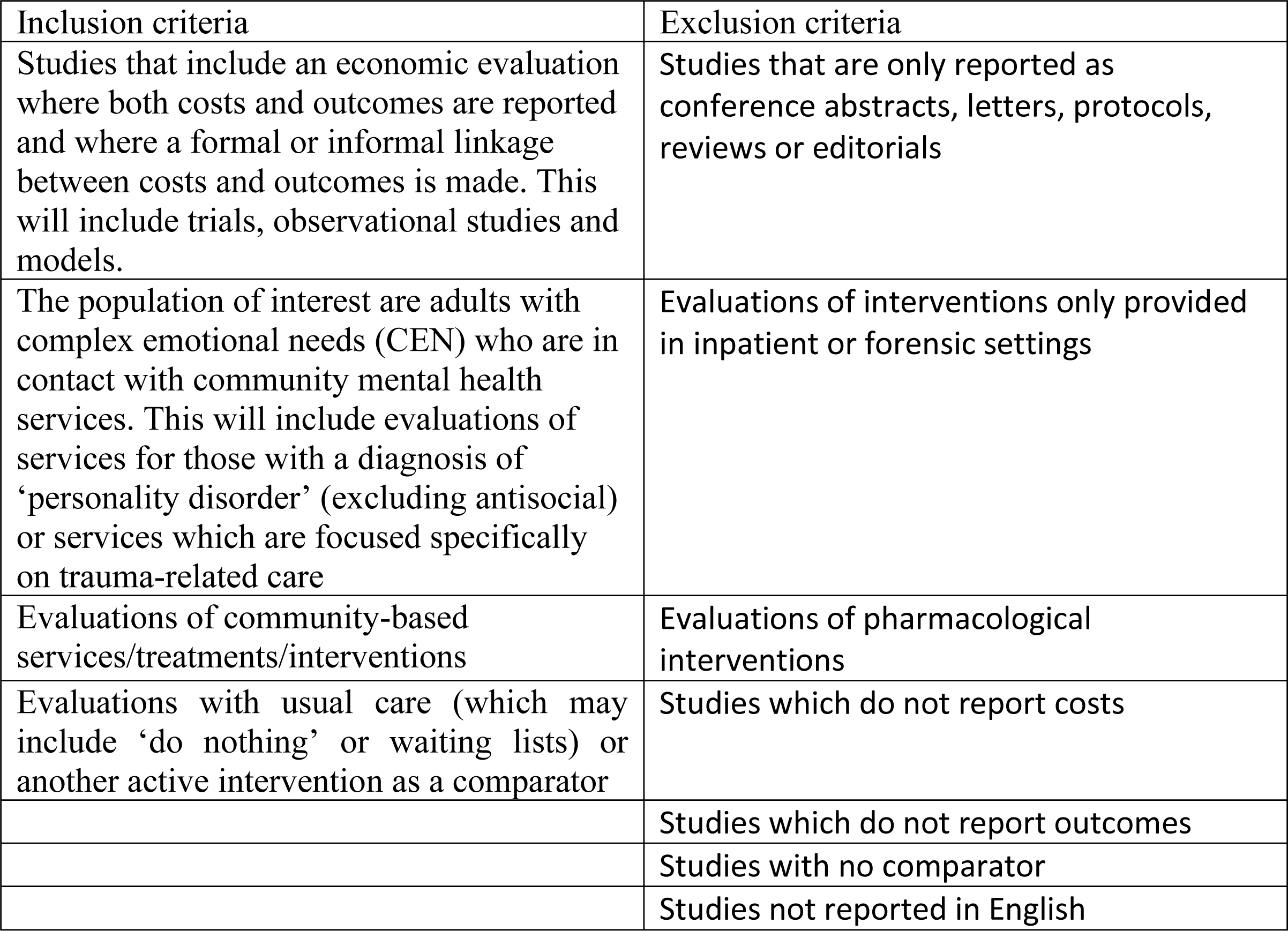
Inclusion and exclusion criteria.

| Inclusion criteria | Exclusion criteria |
| --- | --- |
| Studies that include an economic evaluation where both costs and outcomes are reported and where a formal or informal linkage between costs and outcomes is made. This will include trials, observational studies and models. | Studies that are only reported as conference abstracts, letters, protocols, reviews or editorials |
| The population of interest are adults with complex emotional needs (CEN) who are in contact with community mental health services. This will include evaluations of services for those with a diagnosis of 'personality disorder' (excluding antisocial) or services which are focused specifically on trauma-related care | Evaluations of interventions only provided in inpatient or forensic settings |
| Evaluations of community-based services/treatments/interventions | Evaluations of pharmacological interventions |
| Evaluations with usual care (which may include 'do nothing' or waiting lists) or another active intervention as a comparator | Studies which do not report costs |
|  | Studies which do not report outcomes |
|  | Studies with no comparator |
|  | Studies not reported in English |

Publication date was limited to January 1^st^1990 to September 18^th^2019, and language of publication to English. Studies of interventions for adults with diagnosed ‘antisocial personality disorder’ were excluded, as there are often high levels of forensic service use which would limit or prevent the implementation of community-based interventions. Cost-minimisation studies were excluded as they require interventions to have an equivalent effect, which would be incredibly unlikely in this setting. Community-based services, treatments and interventions are defined as any services, treatments and interventions provided outside of an inpatient setting excluding pharmacological treatments.

### Information sources and search terms

Electronic searches of the following bibliographic databases were conducted: MEDLINE, PsychINFO, EMBASE, Global Health, and NHSEED database. Search terms can be found in the appendix. Search domains covered are: ‘Personality Disorder’, ‘Community Care’ and ‘Economic Evaluation’. Citation tracking of included studies was conducted using Google Scholar. Reference lists of included studies were also screened.

### Selection process and data collection

Three authors (JB, TS and RS) screened titles and abstracts against the inclusion criteria independently using a systematic review web application called Rayyan.qcri.org, which has a blinding feature. A third author (PM) independently reviewed and resolved disagreements. Full text screening was conducted by JB and TS independently (with PM resolving disagreements) using eligibility forms pertaining to each aspect of the inclusion criteria. Reviewers were not blinded to authors, institutions, or journals.

Data were extracted into standard forms by JB. Duplicate extraction was completed by TS for 25% of papers and PM resolved disagreements. Data were collected on the characteristics, methods, and results of each study. The study characteristics data included the country, funder, intervention (including levels of contact), the comparator, study design, economic approach (i.e. whether a cost-effectiveness or cost-utility analysis was used), a summary of the inclusion and exclusion criteria, the sample size, and sample characteristics (mean age, proportion female and proportion employed).

Data on the study methods included the follow-up period, perspective, costing (approach to recording service use, valuing the intervention, sources of unit costs, and discounting), and outcomes (main economic outcome, QALY derivation if relevant, and discounting). The perspective is the range of costs included in an analysis. A narrow perspective would only include costs relevant to the service provider. A broad perspective would include costs to other parts of society, such as the criminal justice system, social services, a patient’s employer, or voluntary/informal care. Inclusion of all impacts would constitute a societal perspective which is rarely achieved. However, here we follow the convention of defining a societal perspective as including healthcare and lost workdays. Resource use measurement methods include reviewing hospital records or conducting patient interviews/surveys. The source of unit costs includes national routine data, information from local providers, and fees or tariffs. The outcome is the measure of effectiveness used in the evaluation. This can be specific to a condition, such as an improvement in a condition-specific scale, or generic such as QALYs. For both costs and outcomes, data on any discounting has been recorded. Discounting is used to reflect the perceived lower value attached to future cost and outcomes, due to the widely accepted view of positive time preference. It is conventional to only discount costs and outcomes that occur beyond a one-year period (9).

### Data analysis

Meta-analysis was not undertaken as the economic evaluations are context specific and the review does not focus on one particular outcome. Narrative synthesis was undertaken to describe the main study findings based on the reporting of the data items described above, including outcomes and costs for the intervention and comparator groups, and the cost-utility or cost effectiveness results. Results were often expressed as incremental cost-effectiveness or cost-utility ratios. These ratios are typically derived from mean values and due to variation around the mean, consideration should be given to uncertainty in these estimates.

### Quality assessment

Study quality was assessed using the Consolidated Health Economic Evaluation Reporting Standards (CHEERS) statement, which sets out a standard for the reporting of economic evaluations (13). The statement evaluates quality of reporting rather than the quality of evidence. Two statements relate to the introduction, sixteen to the methods, five to the results, and there is one statement each for the title, abstract, sources of funding, and conflicts of interest. JB appraised 50% of papers and AC appraised the remaining 50%. TS provided an independent appraisal of 25% of papers and any disagreements were resolved by PM. Each paper will be assigned a score, which corresponds to how many of the 28 statements on the CHEERS checklist the paper complies with (0 = does not comply, 0.5 = partially complies, and 1 = fully complies). Some of the statements in the checklist relate to the quality and fullness of reporting, which can be influenced by publications (e.g. the structure of the abstract) rather than the quality of research itself. A breakdown of each paper’s score has been reported in the results and in table 6, so scores for methodology and results can be assessed independently.

**Table 2.**
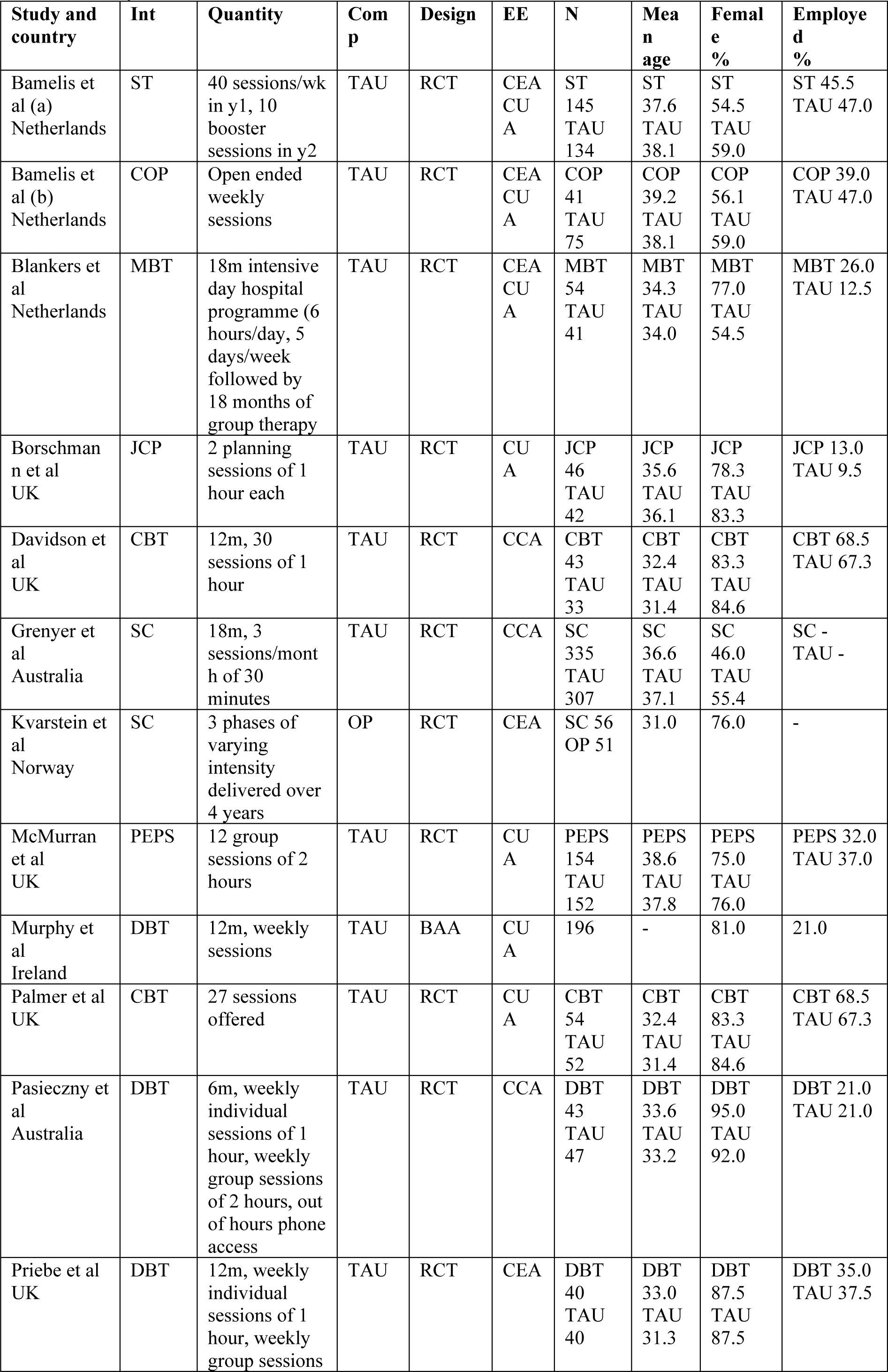
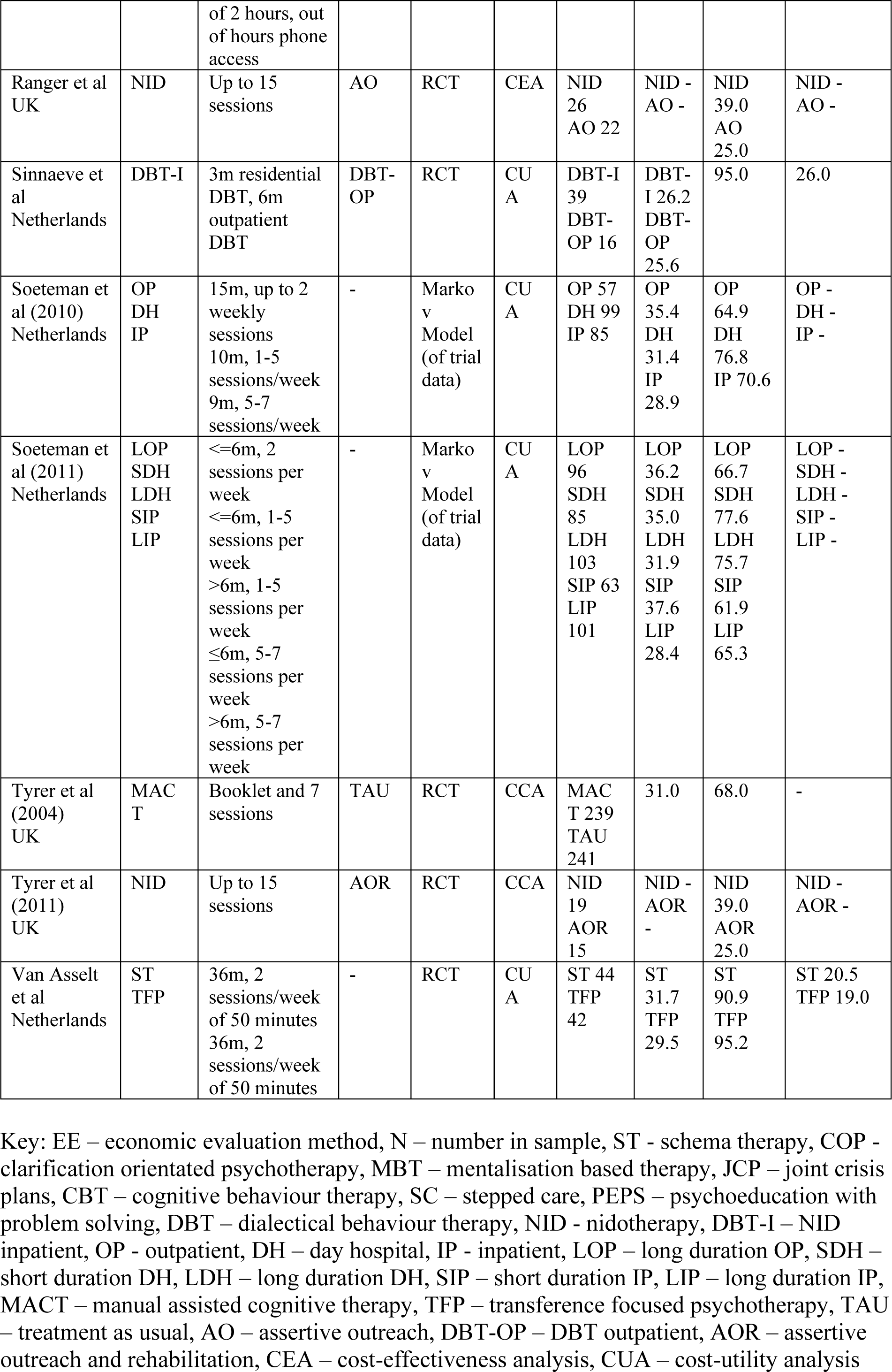
Study characteristics.

| Study and country | Int | Quantity | Comp | Design | EE | N | Mean age | Female % | Employed % |
| --- | --- | --- | --- | --- | --- | --- | --- | --- | --- |
| Bamelis et al (a) Netherlands | ST | 40 sessions/wk in y1, 10 booster sessions in y2 | TAU | RCT | CEA<br>CU<br>A | ST<br>145<br>TAU<br>134 | ST<br>37.6<br>TAU<br>38.1 | ST<br>54.5<br>TAU<br>59.0 | ST 45.5<br>TAU 47.0 |
| Bamelis et al (b) Netherlands | COP | Open ended weekly sessions | TAU | RCT | CEA<br>CU<br>A | COP<br>41<br>TAU<br>75 | COP<br>39.2<br>TAU<br>38.1 | COP<br>56.1<br>TAU<br>59.0 | COP 39.0<br>TAU 47.0 |
| Blankers et al Netherlands | MBT | 18m intensive day hospital programme (6 hours/day, 5 days/week followed by 18 months of group therapy | TAU | RCT | CEA<br>CU<br>A | MBT<br>54<br>TAU<br>41 | MBT<br>34.3<br>TAU<br>34.0 | MBT<br>77.0<br>TAU<br>54.5 | MBT 26.0<br>TAU 12.5 |
| Borschmann et al UK | JCP | 2 planning sessions of 1 hour each | TAU | RCT | CU<br>A | JCP<br>46<br>TAU<br>42 | JCP<br>35.6<br>TAU<br>36.1 | JCP<br>78.3<br>TAU<br>83.3 | JCP 13.0<br>TAU 9.5 |
| Davidson et al UK | CBT | 12m, 30 sessions of 1 hour | TAU | RCT | CCA | CBT<br>43<br>TAU<br>33 | CBT<br>32.4<br>TAU<br>31.4 | CBT<br>83.3<br>TAU<br>84.6 | CBT 68.5<br>TAU 67.3 |
| Grenyer et al Australia | SC | 18m, 3 sessions/month of 30 minutes | TAU | RCT | CCA | SC<br>335<br>TAU<br>307 | SC<br>36.6<br>TAU<br>37.1 | SC<br>46.0<br>TAU<br>55.4 | SC -<br>TAU - |
| Kvarstein et al Norway | SC | 3 phases of varying intensity delivered over 4 years | OP | RCT | CEA | SC 56<br>OP 51 | 31.0 | 76.0 | - |
| McMurren et al UK | PEPS | 12 group sessions of 2 hours | TAU | RCT | CU<br>A | PEPS<br>154<br>TAU<br>152 | PEPS<br>38.6<br>TAU<br>37.8 | PEPS<br>75.0<br>TAU<br>76.0 | PEPS 32.0<br>TAU 37.0 |
| Murphy et al Ireland | DBT | 12m, weekly sessions | TAU | BAA | CU<br>A | 196 | - | 81.0 | 21.0 |
| Palmer et al UK | CBT | 27 sessions offered | TAU | RCT | CU<br>A | CBT<br>54<br>TAU<br>52 | CBT<br>32.4<br>TAU<br>31.4 | CBT<br>83.3<br>TAU<br>84.6 | CBT 68.5<br>TAU 67.3 |
| Pasieczny et al Australia | DBT | 6m, weekly individual sessions of 1 hour, weekly group sessions of 2 hours, out of hours phone access | TAU | RCT | CCA | DBT<br>43<br>TAU<br>47 | DBT<br>33.6<br>TAU<br>33.2 | DBT<br>95.0<br>TAU<br>92.0 | DBT 21.0<br>TAU 21.0 |
| Priebe et al UK | DBT | 12m, weekly individual sessions of 1 hour, weekly group sessions of 2 hours, out of hours phone access | TAU | RCT | CEA | DBT<br>40<br>TAU<br>40 | DBT<br>33.0<br>TAU<br>31.3 | DBT<br>87.5<br>TAU<br>87.5 | DBT 35.0<br>TAU 37.5 |
| Ranger et al UK | NID | Up to 15 sessions | AO | RCT | CEA | NID 26<br>AO 22 | NID - AO - | NID 39.0<br>AO 25.0 | NID - AO - |
| Sinnaeve et al Netherlands | DBT-I | 3m residential DBT, 6m outpatient DBT | DBT-OP | RCT | CUA | DBT-I 39<br>DBT-OP 16 | DBT-I 26.2<br>DBT-OP 25.6 | 95.0 | 26.0 |
| Soeteman et al (2010) Netherlands | OP<br>DH<br>IP | 15m, up to 2 weekly sessions<br>10m, 1-5 sessions/week<br>9m, 5-7 sessions/week | - | Markov Model (of trial data) | CUA | OP 57<br>DH 99<br>IP 85 | OP 35.4<br>DH 31.4<br>IP 28.9 | OP 64.9<br>DH 76.8<br>IP 70.6 | OP - DH - IP - |
| Soeteman et al (2011) Netherlands | LOP<br>SDH<br>LDH<br>SIP<br>LIP | <=6m, 2 sessions per week<br><=6m, 1-5 sessions per week<br>>6m, 1-5 sessions per week<br>≤6m, 5-7 sessions per week<br>>6m, 5-7 sessions per week | - | Markov Model (of trial data) | CUA | LOP 96<br>SDH 85<br>LDH 103<br>SIP 63<br>LIP 101 | LOP 36.2<br>SDH 35.0<br>LDH 31.9<br>SIP 37.6<br>LIP 28.4 | LOP 66.7<br>SDH 77.6<br>LDH 75.7<br>SIP 61.9<br>LIP 65.3 | LOP - SDH - LDH - SIP - LIP - |
| Tyrer et al (2004) UK | MACT | Booklet and 7 sessions | TAU | RCT | CCA | MACT 239<br>TAU 241 | 31.0 | 68.0 | - |
| Tyrer et al (2011) UK | NID | Up to 15 sessions | AOR | RCT | CCA | NID 19<br>AOR 15 | NID - AOR - | NID 39.0<br>AOR 25.0 | NID - AOR - |
| Van Asselt et al Netherlands | ST<br>TFP | 36m, 2 sessions/week of 50 minutes<br>36m, 2 sessions/week of 50 minutes | - | RCT | CUA | ST 44<br>TFP 42 | ST 31.7<br>TFP 29.5 | ST 90.9<br>TFP 95.2 | ST 20.5<br>TFP 19.0 |

Key: EE – economic evaluation method, N – number in sample, ST - schema therapy, COP - clarification orientated psychotherapy, MBT – mentalisation based therapy, JCP – joint crisis plans, CBT – cognitive behaviour therapy, SC – stepped care, PEPS – psychoeducation with problem solving, DBT – dialectical behaviour therapy, NID - nidotherapy, DBT-I – NID inpatient, OP - outpatient, DH – day hospital, IP - inpatient, LOP – long duration OP, SDH – short duration DH, LDH – long duration DH, SIP – short duration IP, LIP – long duration IP, MACT – manual assisted cognitive therapy, TFP – transference focused psychotherapy, TAU – treatment as usual, AO – assertive outreach, DBT-OP – DBT outpatient, AOR – assertive outreach and rehabilitation, CEA – cost-effectiveness analysis, CUA – cost-utility analysis

**Table 3.** Further study characteristics.

| Study and country | Follo w-up period | Costing perspective | Service use measure | Unit costs | Unit costs | Cost discount rate | Main economic outcome | QAL Y measure | QAL Y discount rate |
| --- | --- | --- | --- | --- | --- | --- | --- | --- | --- |
| Bamelis et al (a) Netherlands | 36 | Societal | Combination of patient reported & hospital records | - | National averages | 4% | QALYs & SCID-II. | EQ-5D (UK tariff) | - |
| Bamelis et al (b) Netherlands | 36 | Societal | Combination of patient reported & hospital records | - | National averages | 4% | QALYs & SCID-II. | EQ-5D (UK tariff) | - |
| Blankers et al Netherlands | 36 | Societal | TiC-P | - | national averages | 4% | QALYs (discounted at 1.5%) & BPDSI <15 | EQ-5D (Dutch tariff). | 1.5% |
| Borschmann et al UK | 6 | Health & Social Care | Combination of patient reported - AD-SUS - and hospital records. | Micro-costing approach | National averages. | - | QALYs | EQ-5D (UK tariff) | - |
| Davidson et al UK | 72 | Health, social care & Criminal justice | Combination of patient reported - CSRI - and hospital records. | - | - | - | QALYs | EQ-5D (UK tariff) | - |
| Grenyer et al Australia | 36 | Inpatient costs | Patient/hospital records | - | National average. | - | Frequency & length of inpatient stays. | NA. | - |
| Kvarstein et al Norway | 48 | Health & social care | Patient reported. | Top-down. | National averages. | - | GAF | NA. | - |
| McMurrain et al UK | 72 | Societal | Patient reported - CSRI. | Micro-costing approach | National averages | - | QALYs | EQ-5D (Irish tariff) | - |
| Murphy et al Ireland | 18 | Health service. | Patient reported - 'dedicated resource use questionnaire'. | Micro-costing approach | DRG estimates, literature & national averages | - | QALYs | EQ-5D (Irish tariff) | - |
| Palmer et al UK | 24 | Societal | Combination of patient reported - CSRI - and hospital records. | Micro-costing approach | Local/national data sets | 3.5% | QALYs - discounted at 3.5% | EQ-5D (UK tariff) | 4% |
|  |  |  |  |  |  |  | annually |  |  |
| Pasieczny et al<br>Australia | 6 | Health service | Hospital records. | Micro-costing approach | Local costs. | - | NSSI, ED visits, frequency & length of inpatient stays, BDI-II, BSS, STAI-Y, & BSI | NA. | - |
| Priebe et al<br>UK | 12 | Societal | Patient reported - CSRI. |  | National averages. | - | Percentage point difference in self-harm rates. | NA. | - |
| Ranger et al<br>UK | 12 | Societal | Patient reported - Secure Facilities Use Schedule (SFSUS). | Micro-costing approach | National averages. | - | BPRS | NA. | - |
| Sinnaeve et al<br>Netherlands | 12 | Health service & employment | Patient reported - TiC-P. | - | National averages. | - | QALYs | EQ-5D (Dutch tariff). | - |
| Soeteman et al<br>(2010)<br>Netherlands | 60 | Societal | Combination of patient reported - TiC-P - and hospital records. | Micro-costing approach. | National averages. | - | QALYs & recovered patient years | EQ-5D (Dutch tariff) | - |
| Soeteman et al<br>(2011)<br>Netherlands | 60 | Societal | Combination of patient reported - TiC-P - and hospital records. | Micro-costing approach. | National averages. | 3% | QALYs & recovered patient years | EQ-5D (Dutch tariff) | - |
| Tyrer et al (2004)<br>UK | 12 | Societal | Patient reported - CSRI. | - | National averages. | - | Linehan's Parasuicide History Interview | NA. | - |
| Tyrer et al (2011)<br>UK | 12 | Societal | Patient reported - SFSUS. | Micro-costing approach. | National averages. | - | Bed-days | NA. | - |
| Van Asselt et al | 48 | Societal | Patient reported - interview. | Micro-costing approach. | National averages. | 4% | QALYs | EQ-5D (UK tariff) | 4% |
| Netherla<br>nds |  |  |  |  |  |  |  |  |  |

**Table 4.**
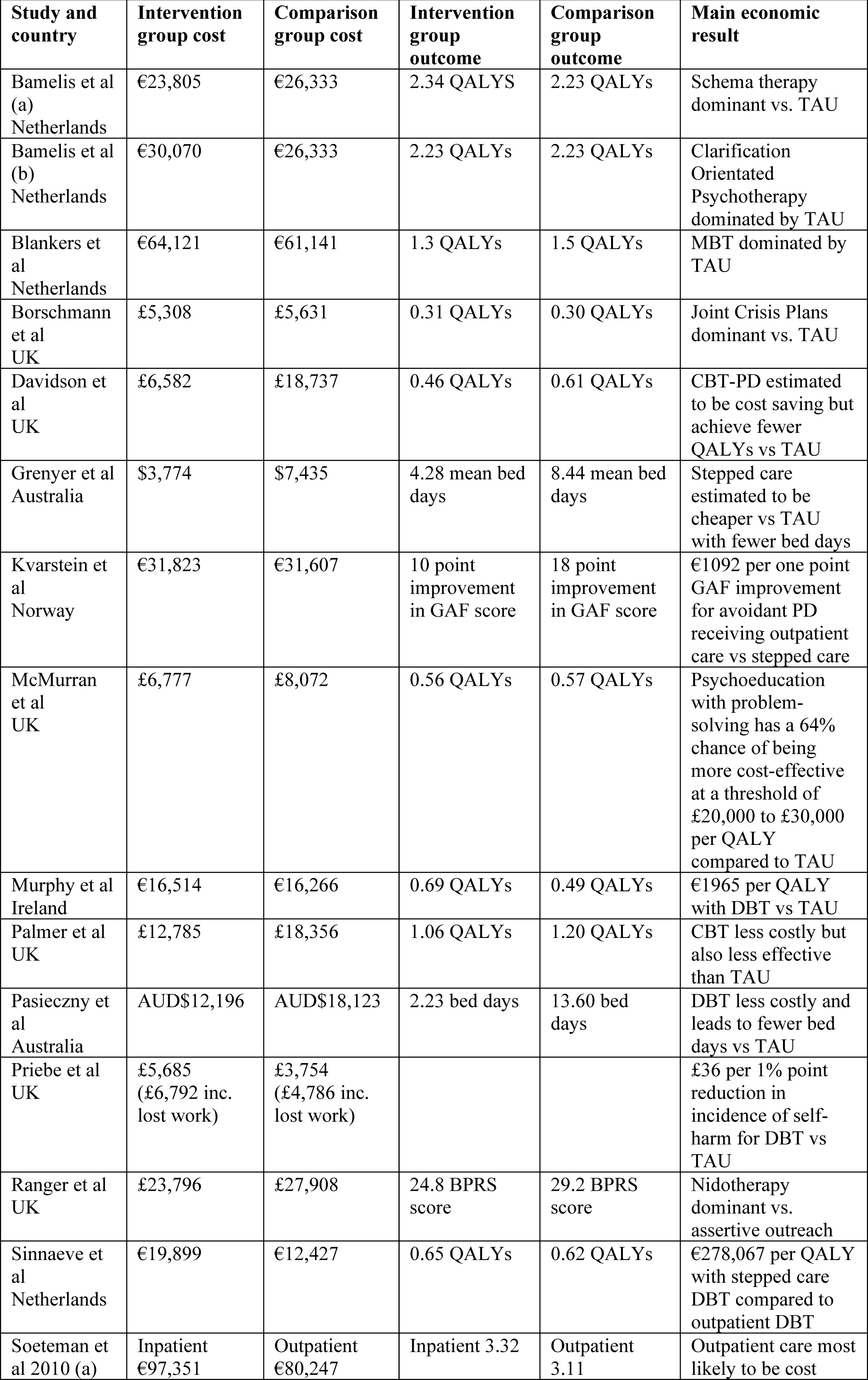
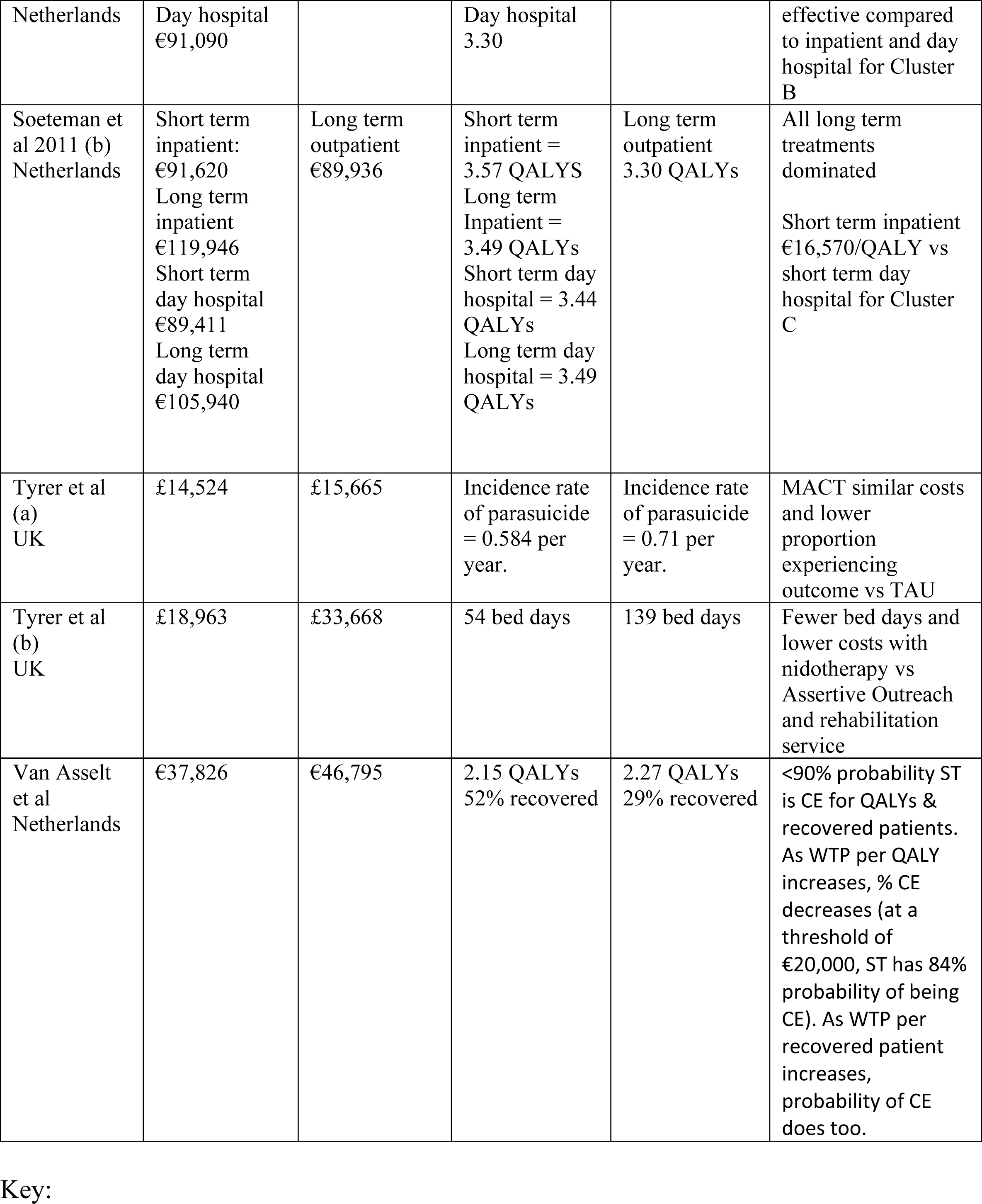
Study results.

**Table 5.** outcomes used in studies.

| *Outcome* | *Measure(s)* | *Papers* | *Prim/Sec* |
| --- | --- | --- | --- |
| Recovered patients | SCID-II | Bamelis et al | Primary |
|  |  | Davidson et al |  |
|  | ADP-IV | Bamelis et al | Primary (back-up)* |
|  | BPDSI | Blankers et al | Effectiveness Measure |
|  |  | van Asselt et al |  |
| HRQoL | EQ-5D-3L | Bamelis et al | Economic outcome |
|  |  | Blankers et al | Utility measure |
|  |  | Borschmann et al | Secondary |
|  |  | McMurrin et al |  |
|  |  | Palmer et al | Primary |
|  |  | Davidson et al |  |
|  |  | Sinnaeve et al |  |
|  |  | Soeteman et al (2010) |  |
|  |  | Soeteman et al (2011) |  |
|  |  | van Asselt et al |  |
|  | EQ-5D-5L | Murphy et al |  |
| Self-harm/NSSI | Self-report questionnaire | Borschmann et al | Primary |
|  | ADSHI | Davidson et al |  |
|  | Structured Interview | Priebe et al | Primary |
|  |  | Pasieczny & Connor | Clinical service measure |
|  | LPC | Sinnaeve et al |  |
|  | LPHI | Tyrer et al (2004) |  |
| Depression & Anxiety | HADS | Borschmann et al | Secondary |
| Working alliance | WAI | Borschmann et al | Secondary |
| Client satisfaction | CSQ | Borschmann et al | Secondary |
| Client Engagement | SES | Borschmann et al | Secondary |
|  | EAS | Tyrer et al (2011) |  |
| Mental Well-being | WEMWS | Borschmann et al | Secondary |
| Social Functioning | WSAS | Borschmann et al | Secondary |
|  | SFQ | Davidson et al |  |
|  |  | Ranger et al | Secondary |
|  |  | Tyrer et al (2011) | Secondary |
|  | IIP-32 | Davidson et al |  |
| Coercion (during hospital visits) | TES | Borschmann et al | Secondary |
| Frequency of inpatient visits | - | Grenyer et al | Primary |
|  |  | Pasieczny & Connor | Clinical service measure |
| Duration of inpatient visits | - | Grenyer et al | Primary |
|  |  | Pasieczny & Connor | Clinical service measure |
|  |  | Ranger et al | Primary |
|  |  | Tyrer et al (2011) | Primary |
| Frequency of ED visits | - | Grenyer et al | Primary |
|  |  | Pasieczny & Connor | Clinical service measure |
| Functioning | GAF | Kvarsein et al |  |
| Depression | BDI | Davidson et al |  |
|  |  | Pasieczny & Connor | Self-report measure |
| Anxiety | STAI | Davidson et al |  |
|  |  | Pasieczny & Connor | Self-report measure |
| Beliefs related to PD | YSQ | Davidson et al |  |
| Suicide attempts | - | Pasieczny & Connor | Clinical service measure |
|  | LPC | Sinnaeve et al |  |
| Behavioural & Service Utilisation | - | Pasieczny & Connor | Clinical service measure |
| Suicidal planning | BSSI | Pasieczny & Connor | Self-report measure |
| Psychiatric Symptoms | BSI | Pasieczny & Connor | Self-report measure |
|  |  | Priebe et al | Secondary |
|  | BPRS | Priebe et al | Secondary |
|  |  | Ranger et al | Secondary |
|  |  | Tyrer et al (2011) | Secondary |
| BPD Symptoms/Severity | ZRS-BPD | Priebe et al | Secondary |
|  | BPDSI | Sinnaeve et al |  |
| Subjective QoL | MSA-QoL | Priebe et al | Secondary |
| Recovered patient years | - | Soeteman et al (2010) |  |
|  |  | Soeteman et al (2011) |  |
| Personality Status | PAS-Q | Tyrer et al (2004) |  |
| *if data for primary outcome was missing, this was used instead |  |  |  |

Key: ADP-IV - Assessment of DSM-IV Personality Disorder Questionnaire, ADSHI - Acts of Deliberate Self Harm, BDI - Beck Depression Inventory, BPRS - Brief Psychiatric Rating Scale, BSI - Brief Symptom Inventory, BSSI - Beck Scale for Suicidal Ideation, CSQ- Client Satisfaction Questionnaire, EAS - Engagement and Assessment Scale, EQ-5D-3L - EuroQuol EQ-5D-3L, EQ-5D-5L - EuroQuol EQ-5D-5L, GAF - Global Assessment of Functioning scale, HADS - Hospital Anxiety and Depression Scale, IIP-32 - Inventory of Interpersonal Problems - Short Form 32, LPC - Life-time Parasuicide Count, LPHI - Linehams Parasuicide History Interview, MSA-QoL- Manchester Short Assessment of Quality of Life, PAS-Q - Quick Personality Assessment Schedule, SCID-II- Structured Clinical Interview for DSM-IV Axis II disorders, SES - Service Engagement Scale, SFQ - Social Functioning Questionnaire, STAI - Spielberger State-trait Anxiety Inventory, TES- Treatment Experience Scale, WAI- Working Alliance Inventory, WEMWS - Warwick-Edinburgh Mental Well-being Scale, WSAS - Work and Social Adjustment Scale, YSQ- Young Schema Questionnaire, ZRS-BPD - Zanari Rating Scale for Borderline Personality Disorder

**Table 6.** Quality Appraisal.

|  | Overall |  |  | Title/Abstract |  | Intro |  | Methods |  | Results |  | Discussion |  | D |
| --- | --- | --- | --- | --- | --- | --- | --- | --- | --- | --- | --- | --- | --- | --- |
|  | Score | out of | % | Score | out of | Score | out of | Score | out of | Score | out of | Score | out of | Score |
| *DBT* |  |  |  |  |  |  |  |  |  |  |  |  |  |  |
| Murphy (2019) | 13 | 19 | 68.4% | 2 | 2 | 2 | 2 | 7 | 10 | 1.5 | 2 | 0.5 | 1 |  |
| Pasieczny (2010) | 9.5 | 19 | 50.0% | 0.5 | 2 | 2 | 2 | 5.5 | 9 | 0.5 | 3 | 1 | 1 |  |
| Priebe (2012) | 15.5 | 19 | 81.6% | 1.5 | 2 | 2 | 2 | 7.5 | 9 | 1.5 | 3 | 1 | 1 |  |
| **Means** | **12.7** | **19.0** | **66.7%** | **1.3** | **2.0** | **2.0** | **2.0** | **6.7** | **9.3** | **1.2** | **2.7** | **0.8** | **1.0** |  |
| **SD** | **2.5** | **0.0** | **13.0%** | **0.6** | **0.0** | **0.0** | **0.0** | **0.8** | **0.5** | **0.5** | **0.5** | **0.2** | **0.0** |  |
| *CBT* |  |  |  |  |  |  |  |  |  |  |  |  |  |  |
| Davidson (2010) | 11 | 19 | 57.9% | 1 | 2 | 2 | 2 | 4 | 10 | 2 | 2 | 1 | 1 |  |
| Palmer (2006) | 17 | 19 | 89.5% | 2 | 2 | 1.5 | 2 | 9.5 | 10 | 2 | 2 | 1 | 1 |  |
| Tyrer (2004) | 13 | 19 | 68.4% | 0.5 | 2 | 1.5 | 2 | 6.5 | 9 | 3 | 3 | 0.5 | 1 |  |
| **Means** | **13.7** | **19.0** | **71.9%** | **1.2** | **2.0** | **1.7** | **2.0** | **6.7** | **9.7** | **2.3** | **2.3** | **0.8** | **1.0** |  |
| **SD** | **2.5** | **0.0** | **13.1%** | **0.6** | **0.0** | **0.2** | **0.0** | **2.2** | **0.5** | **0.5** | **0.5** | **0.2** | **0.0** |  |
| *Stepped care* |  |  |  |  |  |  |  |  |  |  |  |  |  |  |
| Grenyer (2018) | 12.5 | 19 | 65.8% | 1.5 | 2 | 2 | 2 | 4.5 | 9 | 1.5 | 3 | 1 | 1 |  |
| Kvarstein (2013) | 14.5 | 20 | 72.5% | 1.5 | 2 | 1.5 | 2 | 5.5 | 10 | 3 | 3 | 1 | 1 |  |
| Sinnaeve (2018) | 13 | 18 | 72.2% | 1 | 2 | 1.5 | 2 | 6 | 9 | 1.5 | 2 | 1 | 1 |  |
| **Means** | **13.3** | **19.0** | **70.2%** | **1.3** | **2.0** | **1.7** | **2.0** | **5.3** | **9.3** | **2.0** | **2.7** | **1.0** | **1.0** |  |
| **SD** | **0.8** | **0.8** | **3.1%** | **0.2** | **0.0** | **0.2** | **0.0** | **0.6** | **0.5** | **0.7** | **0.5** | **0.0** | **0.0** |  |
| *Nidotherapy* |  |  |  |  |  |  |  |  |  |  |  |  |  |  |
| Ranger (2009) | 12 | 18.5 | 66.7% | 1.5 | 2 | 0.5 | 2 | 8.5 | 9 | 1 | 2 | 0.5 | 1 |  |
| Tyrer (2011) | 14 | 17 | 82.4% | 1 | 2 | 1 | 2 | 8 | 9 | 1 | 1 | 1 | 1 |  |
| **Means** | **13.0** | **17.5** | **74.5%** | **1.3** | **2.0** | **0.8** | **2.0** | **8.3** | **9.0** | **1.0** | **1.5** | **0.8** | **1.0** |  |
| **SD** | **1.0** | **0.5** | **7.8%** | **0.3** | **0.0** | **0.3** | **0.0** | **0.3** | **0.0** | **0.0** | **0.5** | **0.3** | **0.0** |  |
| *Schema Therapy* |  |  |  |  |  |  |  |  |  |  |  |  |  |  |
| Bamelis (2015) | 19 | 20 | 95.0% | 2 | 2 | 2 | 2 | 10.5 | 11 | 1.5 | 2 | 1 | 1 |  |
| van Asselt (2008) | 19 | 20 | 95% | 2 | 2 | 2 | 2 | 10 | 11 | 2 | 2 | 1 | 1 |  |
| **Means** | **19** | **20** | **95%** | **2.0** | **2.0** | **2.0** | **2.0** | **10.3** | **11** | **1.8** | **2.0** | **1.0** | **1.0** |  |
| **SD** | **0.0** | **0.0** | **0.0%** | **0.0** | **0.0** | **0.0** | **0.0** | **0.3** | **0.0** | **0.3** | **0.0** | **0.0** | **0.0** |  |
| *Setting* |  |  |  |  |  |  |  |  |  |  |  |  |  |  |
| Soeteman (2010) | 21 | 24 | 87.5% | 2 | 2 | 2 | 2 | 13.5 | 14 | 2.5 | 3 | 1 | 1 |  |
| Soeteman (2011) | 22 | 24 | 81.7% | 2 | 2 | 2 | 2 | 12.5 | 12 | 2.5 | 3 | 1 | 1 |  |
| **Means** | **21.5** | **24.0** | **89.6%** | **2.0** | **2.0** | **2.0** | **2.0** | **13.0** | **14.0** | **2.5** | **3.0** | **1.0** | **1.0** |  |
| **SD** | **0.5** | **0.0** | **2.1%** | **0.0** | **0.0** | **0.0** | **0.0** | **0.5** | **0.0** | **0.0** | **0.0** | **0.0** | **0.0** |  |
| *Other* |  |  |  |  |  |  |  |  |  |  |  |  |  |  |
| Blankers (2019) | 16.5 | 20 | 82.5% | 2 | 2 | 2 | 2 | 8 | 11 | 1.5 | 2 | 1 | 1 |  |
| Borschmann (2013) | 15 | 18 | 83.3% | 1 | 2 | 2 | 2 | 8 | 9 | 2 | 2 | 1 | 1 |  |
| McMurrans (2016) | 16.5 | 19 | 86.8% | 2 | 2 | 2 | 2 | 8 | 10 | 2 | 2 | 0.5 | 1 |  |
| **Means** | **16.0** | **19.0** | **84.2%** | **1.7** | **2.0** | **2.0** | **2.0** | **8.0** | **10.0** | **1.8** | **2.0** | **0.8** | **1.0** |  |
| **SD** | **0.7** | **0.8** | **1.9%** | **0.5** | **0.0** | **0.0** | **0.0** | **0.0** | **0.8** | **0.2** | **0.0** | **0.2** | **0.0** |  |
| **Total Mean** | 15.2 | 19.5 | 78% | 1.5 | 2.0 | 1.8 | 2.0 | 7.9 | 10.2 | 1.8 | 2.3 | 0.9 | 1.0 |  |
| **Total SD** | 2.7 | 1.2 | 0.1 | 0.5 | 0.0 | 0.4 | 0.0 | 2.0 | 1.0 | 0.6 | 0.6 | 0.3 | 0.0 |  |

## Results

Figure 1 shows the PRISMA flowchart pertaining to the review; eighteen papers were included in the review. One paper (Bamelis et al 2015) presented both cost-effectiveness and cost-utility analyses for each of two separate interventions (14). For reporting purposes, these interventions are treated separately (labelled ‘a’ and ‘b’) although both were compared to the same control group.

**Figure 1.**
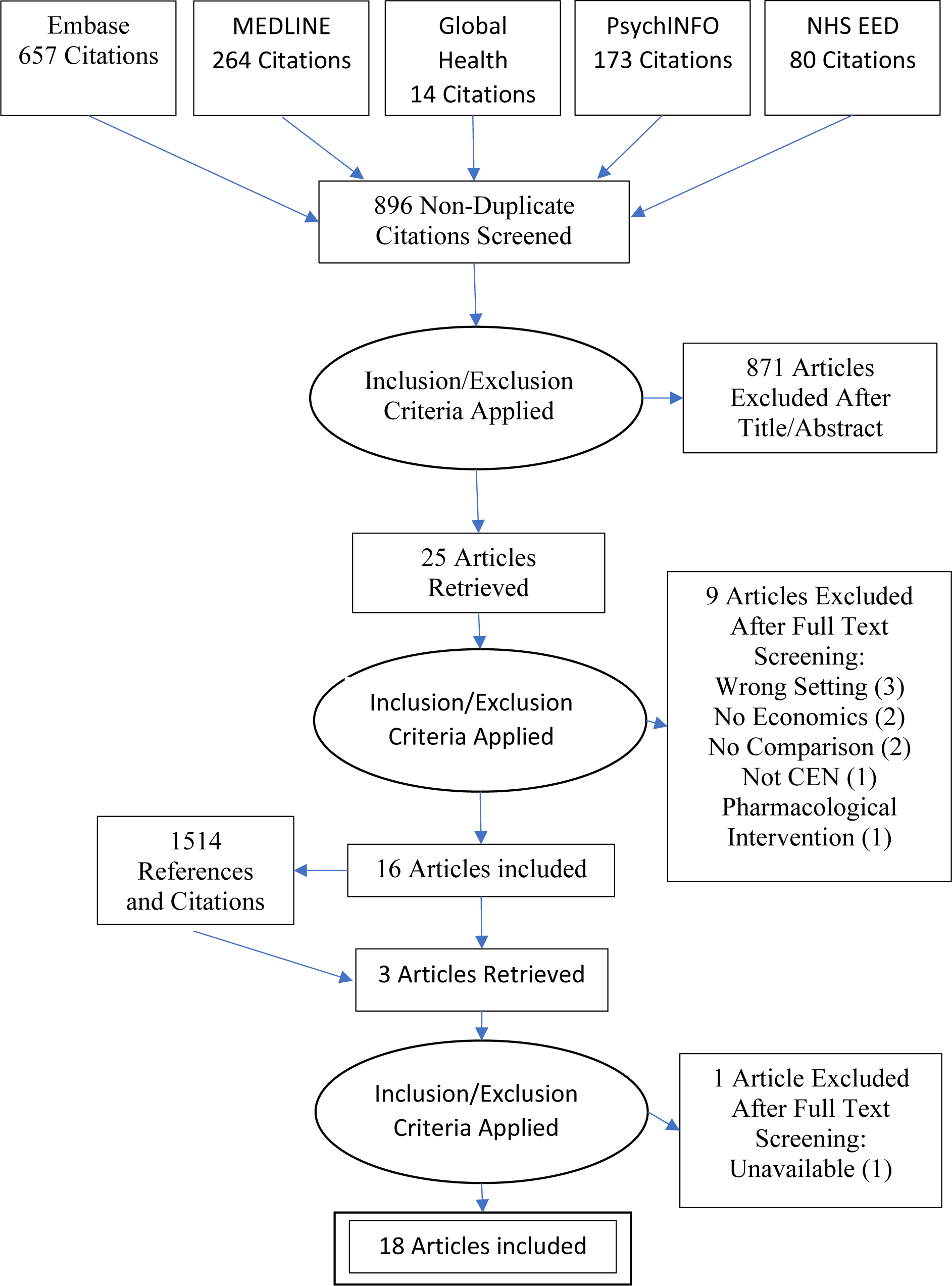
PRISMA flow diagram (12)

PM resolved 14 disagreements between JB & TS during title and abstract screening; two during full text screening; 14 over data extraction; and disagreements in 4 papers in the quality assessment.

### Study Characteristics

Table 2 shows the characteristics of the studies included in the review. Studies were conducted in UK, Europe, and Australia, with sample sizes ranging from 34 to 642 (mean 195.2, SD 167.7). The youngest sample had a mean age of 28.4 years and the oldest sample mean was 39.2 years. The percentage of women in the samples ranged from 25% to 95%. The sample with the lowest rate of employment was 9.5% and the highest rate of employment was 68.5%, though seven studies did not report this.

Five of the studies included patients with any ‘personality disorder’ diagnosis (Grenyer et al 2018, McMurran et al 2016, Kvarstein et al 2013, Priebe et al 2012, Tyrer et al 2011) (15–19). Three of the evaluations looked at ‘Cluster C personality disorders’, described as avoidant, dependent, obsessive-compulsive (Bamelis et al 2015a, Bamelis et al 2015b, Soeteman et al 2011) (14,20) and one study looked at ‘Cluster B personality disorders’ described as antisocial, borderline, histrionic, and narcissistic (Soeteman et al 2010) (21). Eight studies only recruited participants with a ‘borderline personality disorder’ diagnosis (Blankers et al 2019, Borschmann et al 2013, Davidson et al 2010, Palmer et al 2006, Pasieczny & Connor 2011, Sinnaeve et al 2018, van Asselt et al 2008) (22–28). Nine of the studies required a recent episode of self-harm, attendance at an emergency department, or in-patient stay (Borschmann et al 2013, Davidson et al 2010, Grenyer et al 2018, Murphy et al 2019, Palmer et al 2006, Pasieczny & Connor 2011, Priebe et al 2012, Sinnaeve et al 2018, Tyrer et al 2004) (15,18,23–27,29,30). Two studies only included patients with other comorbid severe mental illness (Ranger et al 2009, Tyrer et al 2011) (31,32), of which one of these studies also required substance dependence (Tyrer et al 2011). One study included a general sample of which less than half had complex emotional needs that met personality disorder diagnostic criteria (Tyrer et al 2004) (30).

The interventions evaluated in the 18 studies are described in Table 7. Psychotherapeutic interventions included dialectical behavioural therapy (n=3), types of cognitive therapy (n=3), Nidotherapy (n=2), schema-focused therapy (SFT) (n=2), psycho-education with problem solving (PEPS) (n = 1), and mentalisation based therapy delivered in a day hospital setting (MBT) (n = 1). Other interventions comprised of altering the setting in which care was delivered (n=2), adopting a stepped-care approach (n=3), and developing joint crisis plans (JCPs) (n = 1).

**Table 7:** interventions included in studies.

| **Intervention** | **Description\*\*\*** | **Number of studies identified** | **Study authors** |
| --- | --- | --- | --- |
| **Dialectical behavioural therapy** | A form of psychotherapy adapted from cognitive behavioural therapy. Includes individual therapy, group skills training, phone coaching and consultation meetings for clinicians. | 3 | Murphy et al<br>Pasieczny & Connor<br>Priebe et al |
| **Cognitive behavioural therapy** | Structured, time limited, psycho-social intervention focussing on practical goals to address problems with social functioning. | 2 | Palmer et al<br>Davidson et al |
| **Manual-assisted cognitive therapy** | Brief focused therapy for people with repeated incidents of self-harm. Service users receive a 70-page booklet and are offered up to seven sessions with a therapist which focus on methods to reduce distress and resolve problems. | 1 | Tyrer et al (2004) |
| **Clarification orientated psychotherapy** | Open ended client-centred psychotherapy addressing dysfunctional interaction behaviours delivered via individual outpatient sessions. | 1 | Barnelis et al (2015b)* |
| **Nidotherapy** | Systematic assessment and modification of a service user's physical, social and personal environment through agreed set of targets. | 2 | Ranger et al<br>Tyrer et al (2011) |
| **Schema-focused therapy** | Integrative cognitive therapy combining experiential, behavioural and interpersonal therapy. | 2 | Barnelis et al (2015a)*<br>Van Asselt et al** |
| **Transference-focused psychtherapy** | Psychodynamically based psychotherapy. | 1 | Van Asselt et al** |
| **Psycho-education with problem solving** | Up to four psycho-education sessions discussing diagnosis of 'personality disorder' to improve knowledge. Build rapport and motivate participants. Problem-solving therapy delivered via 12 group sessions aiming to help people learn a strategy for solving interpersonal problems. | 1 | McMurran et al |
| **Mentalisation based therapy in a day hospital setting** | Intensive day hospitalisation for minimum of 18 months including daily group psychotherapy, weekly individual psychotherapy and individual crisis planning alongside social and community meetings and other activities such as art therapy. Followed by 18 months group therapy. | 1 | Blankers et al |
| **Interventions defined by setting** | Out-patient, day hospital and in-patient psychotherapy or psychosocial services. Duration of treatment also explored. | 2 | Soeteman et al 2010<br>Soeteman et al 2011 |
| **Stepped-care approach** | Intensive day, residential or outpatient intervention followed by step down to longer term outpatient/community follow-up psychotherapy. | 3 | Sinnaeve et al<br>Kvarstein et al<br>Grenyer et al |
| **Joint crisis plans** | Written documentation of treatment preferences for management of future crises. Developed between service user and clinician with facilitation from independent mental health professional. | 1 | Borschmann et al |

\*refer to the same study
\*\*refer to the same study
\*\*\*description summarised from studies themselves or relevant corresponding efficacy study

Treatment as usual was the comparative intervention in half of the included papers (n=8). Other comparators included psychotherapy provided by out-patient services (n=4), psychotherapy provided by day-hospital services (n=2), care provided by assertive outreach services (n=2), transference-focused psychotherapy (n=1), and baseline data (n=1). More than three-quarters of the included papers employed a randomised controlled trial design (n= 14), two used Markov models, one a wait-list controlled trial, and one a quasi-experimental design.

A total of 33 different outcome measures were used across the eighteen studies (Table 5) with inconsistent approaches to discounting outcomes (five studies applied a discount rate of between 3-5% and 13 studies applied no discounting). Over half of the papers employed a cost-utility analysis (n=10). Other approaches used were cost-consequences analyses (n=5) and cost-effectiveness analyses (n=5). Outcomes and costs reported by all studies are reported in Table 4.

### DBT

Each of the three studies of DBT used a different method of economic evaluation (Table 2). Murphy et al conducted a quasi-experimental study using a CUA, Pasieczny & Connor adopted a CCA for their wait-list controlled trial, and Priebe et al performed a randomised control trial based CEA(18,26,29). The mean (SD) follow-up period for these studies was 12 (6) months. Murphy et al and Pasieczny & Connor adopted a narrow healthcare perspective, while Priebe et al took a broader societal view including lost employment costs.

Two of the studies (Pasieczny & Connor and Priebe et al) reported significant improvements in clinical outcomes (Table 4). Pasieczny & Connor reported fewer suicide attempts, Emergency Department (ED) visits, admissions and inpatient days. Priebe et al reported a reduction in self-harm events (rate ratio 0.78 (95% confidence interval (CI) 0.76-0.80). Pasieczny & Connor and Murphy et al, both reported DBT to be less costly than the control, though neither reported statistical significance. Murphy et al reported an ICER for DBT of €1965 per QALY, with the sensitivity analysis suggesting a 62% probability that DBT was cost-effective at a threshold (i.e. how much society is considered to value a QALY) of €45,000 EUR per QALY. Priebe et al reported the ICER of DBT was £36 per percentage point decrease in the incidence of self-harm.

Quality appraisal found the mean score of studies evaluating DBT to meet 67% (SD = 13%) of the applicable criteria (Table 5). Priebe et al met the most criteria, scoring 15.5 out of 19 applicable statements. Both Priebe et al and Murphy et al had good scores for the reporting of their methods (7.5 out of 9 and 7 out of 10, respectively) and results (1.5 out of 3 and 1.5 out of 2, respectively). Pasieczny & Connor had the lowest quality scores overall (9.5 of 19).

### Cognitive therapies

Details of the three studies evaluating cognitive therapies are given in Table 3. All three studies used data from randomised controlled trials. Palmer et al carried out a CUA, whereas Davidson et al and Tyrer et al (2004) employed a CCA (24,25,30). Tyrer et al (2004) evaluated manual-assisted cognitive therapy (MACT), with the other two evaluating cognitive behavioural therapy (CBT). The follow-up periods for these studies were the most varied for a specific intervention category (mean 36 months SD 31.75). Tyrer et al (2004) and Palmer et al took a societal perspective and Davidson et al employed a Health, social care and criminal justice perspective. All three used a version of the Client Service Receipt Inventory to record resource use, though Davidson et al and Palmer et al also used hospital records.

Only Tyrer et al (2004) reported a change in outcome that favoured cognitive therapies, the incidence rate of a first episode of self-harm per person year was 0.584 in the MACT group and 0.71 in the TAU group. The frequency of self-harm events remained relatively unchanged (MACT = 2.84 per year vs TAU = 2.54, after outliers had been excluded). Davidson et al and Palmer et al reported the difference in costs between CBT and TAU, though they were not found to be significant. Palmer et al found CBT was less costly but also less effective and reported an ICER of £6376 per QALY. When WTP is below the ICER, CBT has a higher probability of being cost-effective, but when WTP is above the ICER, TAU has a higher probability of being cost-effective. At the UK threshold of £30,000 per QALY, the probability of CBT being cost effective was only ∼25%, whereas the probability of TAU being cost-effective was ∼75%, making TAU the more cost-effective choice. Neither Davidson et al nor Tyrer et al (2004) calculated an ICER.

Quality appraisal found the papers had a mean score of 72% (SD 13%). Palmer et al had the second highest quality appraisal score, scoring 18 points across 19 applicable criteria (95%). Tyrer et al (2004) scored 13 out of 19, failing to meet the title and abstract, and methodological criteria well (0.5 of 2, and 6.5 of 9, respectively). Davidson et al had the lowest QA score of the three, again, scoring poorly on criteria for the title and abstract, and the methods, but also the disclosure criteria (1 of 2, 4 of 10, and 1 of 2, respectively).

### Stepped care

Details of methods used in the three studies of stepped-care interventions are presented in Table 3. All three studies used data from randomised controlled trials. Sinnaeve et al carried out a CUA, Kvarstein et al employed a CEA and Grenyer et al conducted a CCA(15,17,27). The mean (SD) follow up period was 32 (18.33) months. The perspective of Kvarstein et al and Sinnaeve et al included healthcare and one other sector’s costs (social care and employment respectively), while Grenyer et al took the narrowest perspective of all the studies included in the review, only including in-patient stays.

The model of stepped care varied between studies. Grenyer et al considered a brief intervention clinic delivered within 36 hours of a crisis presentation, followed by longer term community based psychological therapy. A significant reduction in inpatient days was reported. Sinnaeve et al evaluated three months of DBT delivered as a residential intervention, followed by six months of outpatient DBT, and compared this to 12 months of standard outpatient DBT. Sinnaeve et al reported QALYs with mean (SD) utility scores; these were higher for the step down DBT care group than the comparator however this difference was not statistically significant (stepped care DBT = 0.65 (0.33) vs outpatient DBT = 0.62 (0.28)). Kvarstein et al evaluated intensive psychotherapy in day hospitals followed by outpatient individual and group therapy. Lower improvements in functioning were reported (measured on the Global Assessment of Functioning scale) compared to standard out-patient care (Table 4).

None of the studies reported significant differences in costs. Sinnaeve et al reported an ICER of €278,067 per QALY;at a threshold of €80,000 per QALY there was a 21% likelihood that stepped care was the most cost-effective option. Kvarstein et al reported an ICER for outpatient care compared to stepped care for the ‘avoidant personality disorder’ subgroup (but not the borderline subgroup) which was €1,092 per additional point gained in the Global Assessment of Functioning scale (GAF) (favouring outpatient care). No uncertainty analysis was reported.

Mean (SD) scores of applicable criteria met during quality appraisal were 70% (3%). Stepped care was the only intervention for which none of the papers scored above 80% on the quality appraisal. All three papers scored well in the criteria relating to the title and abstract, introduction, and discussion. However, all three papers failed to meet multiple criteria for the methods, resulting in a mean score of 5.3 out of 9.3 applicable criteria.

### Nidotherapy

Two studies reported economic evaluations of Nidotherapy using data from a single pilot randomised controlled trial in a population with difficult to manage needs, ‘personality disorder’ diagnoses and other comorbid severe mental illness. Ranger et al evaluated the pilot using a CEA (31). Tyrer et al (2011) later evaluated Nidotherapy in a sub-group of patients with substance misuse issues, employing a CCA using data from the same trial (32). The studies used a broad perspective and a follow-up period of 12 months.

Ranger et al did not observe any significant effects on outcomes, with a trend towards reduced symptoms in the intervention group. Tyrer et al (2011) reported a significant reduction in bed days in secondary sub-group analyses and also found Nidotherapy was less costly, although the difference was not statistically significant at the p<0.05 confidence level. Ranger et al still found Nidotherapy to be dominant (i.e. it resulted in lower costs and better secondary outcomes than the comparator, although these results were not significant) and even at a threshold of £0 per point of improvement in the brief psychiatric rating scale (BPRS) it had a 60% likelihood of being cost-effective.

Quality appraisal scores were 64.9% for Ranger et al and 82% for Tyrer et al (2011). Both studies scored similarly on the reporting of their methods and results, however Ranger et al did not fully meet the quality criteria relating to the introduction, discussion, and disclosure.

### Schema therapy

Two studies evaluated schema therapy using randomised controlled trials, and both conducted a CUA and CEA(14,28). Bamelis et al (2015a) compared schema therapy to TAU and van Asselt et al compared schema therapy to transference-focused psychotherapy. Both studies used the same measure of effectiveness, the proportion of recovered patients, and report this to be higher in the schema therapy group than the comparator group. For Bamelis et al (2015a) this was 81.4% vs 51.2%, respectively (‘recovered’ defined as a SCID-II score ≤15) and van Asselt et al reported this to be 52% vs 29%, respectively (‘recovered’ defined as a BPDSI score ≤15). Bamelis et al (2015a) found schema therapy produced a greater median gain in number of QALYs than TAU, though this was not significant (2.34 vs 2.23; p = 0.51). van Asselt et al report total QALYs for the schema therapy group was lower than transference-focused psychotherapy, this difference was also not significant (2.15 vs 2.27, respectively: 95% confidence interval −0.51 to 0.28). van Asselt et al reported an incremental difference in mean costs: schema therapy was €8,969 less costly than transference-focused psychotherapy, though this difference was not significant (95% CI = −21,775 to 3,546). Bamelis et al (2015a) also reported a lower mean (95% CI) cost for schema therapy compared to TAU: €23,805.00 (21,014 to 26,791) vs €26,333.00 (22,384 to 30,605).

Both studies found that schema therapy was dominant, with regard to cost per recovered patient and cost per QALY. Bamelis et al (2015a) showed schema therapy to have an 80% probability of being cost-effective at a WTP threshold of 0, for both cost per recovered patient and cost per QALY. Van Asselt et al reported the likelihood of schema therapy being cost effective, in terms of recovered patients and QALYs, was over 90% when WTP = 0. Both Bamelis et al (2015a) and van Asselt et al observed that as the threshold for recovered patients increased, the probability of schema therapy being cost-effective also increased, however, as the threshold for QALYs increased, the likelihood of schema therapy being cost-effective decreased.

In the quality appraisal, schema therapy studies scored the highest out of all the interventions, with a mean (SD) of 95% (0.0%) of the applicable criteria met. This was the highest mean quality appraisal score for an intervention (5.4% higher than ‘Setting’ at 89.6%). van Asselt et al fully met all applicable criteria for all sections except the methods, for which the score was 9.5 out of 10. Bamelis et al scored similarly well (19 out of 20) but lost half a point in both the methods and results sections.

### Interventions defined by setting

Two evaluations conducted by Soeteman et al employed Markov models to evaluate the effect of care being provided in different settings for samples with either cluster B or cluster C ‘personality disorder’ diagnoses(20,21), using data from the SCEPTRE trial (33,34). Markov models were used in these cost-utility analyses comparing out-patient, day-hospital and inpatient care (one of the studies varied the duration of day-hospital and inpatient care between long and short term, Soeteman et al 2011). Both studies adopted a broad societal perspective, as well as repeating the models taking a narrower, health service provider perspective.

Soeteman et al reported that for a cluster B ‘personality disorder’ group care at a day-hospital was associated with the greatest QALY gains, and for cluster C ‘personality disorder’ group, short-term inpatient care produced the greatest estimated number of QALYs. For cluster B patients, out-patient care was the least costly and also dominated other settings (being less costly and more effective), and for cluster C patients, short-term day hospital treatment was the least costly, and all long-term options were dominated. Estimated cost effectiveness acceptability curves (CEAC) for cluster B showed out-patient care was 84% likely to be cost-effective when the threshold was €0 per QALY. For cluster C patients the estimated CEAC showed short-term inpatient care to be 60% likely to be cost-effective at a high WTP threshold of €80,000 per QALY compared to short-term day-hospital care.

The mean (SD) quality appraisal score for these studies was 9.6% (2.1%). Both papers scored similarly, fully meeting the title and abstract, introduction, and discussion criteria. As the papers were both models, they had the most applicable methods and results criteria, and scored reasonably well, Soetman et al (2010) scored 13.5 out of 14, and Soetman et al (2011) scored 12.5 out of 14. Both papers lost half a point in the results section for only partially meeting one of the criteria (2.5 out of 3). Soetman et al (2010) did not provide information on funding or conflict of interests, and so scored 0 out of 2 on the disclosure statements.

### Other interventions

Four studies were identified which looked at other interventions: joint crisis plans (JCPs) (23), psycho-education with problem solving (PEPS) (16), clarification orientated psychotherapy (COP) (14) and mentalization-based treatment delivered in a day hospital (MBT) (22). All of them used randomised controlled trial data and employed a CUA, and two of these studies, Blankers et al and Bamelis et al (2015b), also conducted a CEA (Table 2). All of these studies took a societal perspective.

Borschmann et al did not report a significant difference in costs or outcomes however their economic analysis found JCPs to be dominant over usual care, with over 80% probability of being cost-effective when the threshold was £0 per QALY. This is not unusual as even in the absence of a significant clinical effect, there can still be a finding of cost-effectiveness. The latter combines point estimates of cost and outcome differences and in some cases, as here, can show high probabilities of interventions being cost-effective. McMurran et al found PEPs was also dominant, having a 64% chance of being more cost-effective compared to usual care with a threshold of £20,000 to £30,000 per QALY.

Bamelis et al (2015b) found COP was inferior to TAU and SFT. Blankers et al found MBT was dominated by TAU in the CUA but also found that in the CEA the ICER per patient in remission was €29,314. At a threshold of €45,000 the chance of MBT being cost effective (when considering ‘remission’ as an outcome) was only slight (55%).

Quality appraisal of Bamelis et al has already been discussed under the schema therapy results. Mean (SD) quality appraisal scores were the highest for the remaining three studies, being 89% (7.3%). Blankers et al met 83% of applicable criteria in the quality appraisal, meeting fully the criteria for title and abstract, introduction, discussion, and disclosure. In the results they met all but one criterion, which they partially fulfilled (results = 1.5 out 2), and in the methods they failed to meet three of 11 applicable criteria (methods = 8 out of 11). Quality appraisal of Borschmann et al found they met 83% of applicable criteria (15 out of 18). They fully met the criteria set out for the introduction, results and discussion, but were short one point in the title and abstract, methods and disclosure sections. McMurran et al met 16.5 of a possible 19 (87%) during quality appraisal. They fully met the criteria for the title and abstract, introduction, results, and disclosure. However, they failed to meet two criteria in the methods (score = 8 out of 10) and only partially met the criteria for the discussion (0.5 out of 1).

## Discussion

### Summary of main findings

This paper has reviewed economic evaluations of community-based interventions for people with complex emotional needs meeting criteria for ‘personality disorder’ diagnoses. A diverse range of interventions were identified, with no strong evidence found for the cost-effectiveness of any single intervention or model of care.

The strongest evidence was for DBT delivered in community settings: all three identified studies indicated the intervention is likely to be cost-effective compared to treatment as usual. However, consideration should be given to the limited pool of economic evidence when interpreting this finding. The review also identified evidence to support the use of schema focused therapy, joint crisis plans, stepped care (as described by Grenyer et al), Nidotherapy, psychoeducation with problem-solving, and MACT. The authors with lived experience highlighted that the review did not identify any economic evaluations which considered service user led interventions or workforce development interventions, such as those which focus on therapeutic alliance.

Across all 18 studies the evidence was weakened by small sample sizes or quality of evidence. Of the 12 studies which found evidence to favour the intervention evaluated, only five reported a statistically significant effect (however, even non-significant effects when combined with cost differences can indicate cost-effectiveness). Of the five studies with significant effects, there were also limitations around reliability of evidence in at least three of the studies: Grenyer et al took a narrow health provider perspective to evaluate their stepped care intervention, limiting relevance for health policy makers (15). The drivers for observed differences between groups were unclear and although significant differences in bed days were observed, there was no difference in admission rates overall (however, reduced bed days alone may be an important effect). The authors also acknowledge that as there were staff transfers between sites in the cluster RCT design, the reliability of evidence may be limited. Pasieczny & Connor used a non-randomised trial design which can lead to biased estimates of effect (26). Tyrer et al (2011) relied on a subsample of trial participants; unplanned subgroup analysis of trial data can lead to unreliable results by increasing the risk of chance findings (32).

### Quality of evidence

While DBT, CBT and stepped care have been the most extensively researched, the numbers of economic evaluations for each of these interventions are relatively few and provide insufficient evidence upon which decision makers can confidently base guidelines or allocate resources. This contrasts with other areas such as depression or schizophrenia where reasonable agreement about interventions exist. The review found interventions were sometimes poorly described, limiting reproducibility and usefulness for implementation decisions. Several studies used data from the same trials to report subsequent sub-group analysis or longer term follow up (BOSCOT/POMCAT/SCEPTRE). This approach weakens evidence as risk of chance findings are increased and bias may be repeated across more than one study.

### Limitations

Our search strategy included ‘community/outpatient’ terms which may have excluded some relevant studies which did not mention the setting in the title or abstract or where care was delivered in a day service. The potential impact of this is likely to be reduced by our supplementary strategies of citation tracking and reference list screening. As has been found in two related reviews(3,4), there was significant heterogeneity in the included studies which prevented meta-analysis of findings. The results are therefore reported in narrative form making interpretation of findings more complex. In addition, the comparator was most often ‘usual care’ which is context specific and rarely described in detail limiting generalisability. The review scope did not intend to include interventions for people with ‘antisocial personality disorder’ diagnoses. However, some of the identified studies considered samples with ‘any personality disorder diagnosis’, another required that patients had other comorbid severe mental illness, and another sample included individuals with complex emotional needs meeting ‘personality disorder’ diagnostic criteria as well as those not meeting this criteria. Given the general nature of the samples, and because these studies met all other inclusion criteria, they were included in the review. However, this presents a key limitation for identifying and interpreting findings specific to the population on interest and highlights that diagnostic categorisation, whilst essential for literature searching, is a limitation of the review scope. Finally, to simplify reporting, we assigned a quality assessment score to each paper which may risk over simplifying quality assessment where it is important to understand the areas of strength and weakness. To address this limitation, we have provided detail on areas of quality assessed in Table 6.

### Recommendations for future research

The majority of included studies relied on data from small randomised controlled trials (RCTs). Whilst RCTs are the gold standard of evidence generation on the effectiveness of treatment, efficacy evidence can also be generated through evaluations in ‘real world’ settings. Such evidence can be more informative for decision making as results are often more generalisable. Economic modelling can also support decision making whilst presenting uncertainty in choices. Given the scarcity of economic evidence in this area, observational studies alongside the delivery of community services would be valuable.

The review found that while a range of economic approaches were used, studies using CUAs applied the most rigorous and transferable methods. The follow-up periods were longer, the perspectives were broader, and resource use was more often collected through a combination of patient report and patient records improving the accuracy of healthcare utilisation estimates. The main benefit of CUAs is that they enable comparison across disease areas by measuring effect in QALYs. All CUAs in this review used the EQ-5D to derive QALYs (in line with NICE guidelines). The EQ-5D has been criticised for not picking up all important aspects of health, and so for all health conditions consideration must be given to its reliability (does it produce consistent results), validity (does it measure what it intends to measure), and sensitivity (does the instrument identify genuine changes in health states). The EQ-5D has been shown to be moderately responsive to change in symptoms in individuals with ‘personality disorder’ diagnoses (35). There is a lack of evidence on its validity in capturing all important aspects of health in this population (36). Nonetheless, due to the measure’s simplicity and because it can be used to generate QALYs, it has been recommended as appropriate for use in this population (35).

A limitation of CUAs is that QALYs singularly focus on health benefits which prevents comparisons across sectors (e.g. comparing whether an education intervention may be more cost-effective than a health intervention) and may underestimate benefits where interventions have wider outcomes such as employment. A majority of studies included in this review took a societal perspective in measuring costs and effects. Five studies also used CCAs, presenting costs alongside a number of outcomes. Whilst this can make interpretation challenging, CCAs may be justified given the multifaceted nature of the conditions being studied and the potential breadth of effects. A broad perspective and presenting multiple outcomes alongside QALYs may therefore be appropriate where interventions aim to improve outcomes beyond health gain, and where cost implications may fall outside of the health and care budget.

Future research should aim to co-produce studies with people with lived experience of diagnoses of ‘personality disorder’ or complex emotional needs to help ensure important outcomes, as well as costs, are captured from a broad perspective. Choice of outcome measures should also be informed by previous studies and local guidelines. Consistency in measurement and reporting will support the development of a stronger evidence base for community-based interventions as information can be pooled across multiple studies. Researchers may wish to consult the ICHOM Standard Set for Personality Disorders when selecting measures (37).

Studies must only evaluate therapies which are well developed, hold face validity with people who are using and delivering services, and must be based on sound theoretical foundations. High quality research is also needed on models of care, including the intensity and duration of interventions, with clear description of how services are configured to enable reproducibility. Researchers may wish to refer to the forthcoming Finamore et al. “Systematic scoping of community-based service models for people with personality disorder” to support a standardised description and understanding of models of care. The logic models articulated in this paper may also support the identification of relevant outcome measures. Finally, the review identified a gap in the type of interventions evaluated with no service user led interventions or workforce interventions identified. These two areas should be considered key areas for future economic research.

### Lived Experience Commentary by Eva Broeckelmann

Having spent years struggling to access suitable treatment for CEN, I am pleased to see the lack of robust economic evidence to support a *single* intervention or model of community-based care.

Especially considering the inherently heterogeneous nature of any given sample with a “PD” diagnosis - where e.g. two people with “BPD” may have no more than one highly subjective trait in common -, there simply is no ‘one-size-fits-all’ approach. Therefore, any study results must be treated with utmost caution before making generic policy decisions that indiscriminately apply to everyone with this label.

Despite being recommended by NICE, I do not consider the EQ-5D to be an appropriate outcome measure for this population. CEN symptoms can fluctuate so frequently that any snapshot in time on such a rudimentary measure is virtually meaningless for assessing long-term recovery, and it will be crucial to develop more nuanced alternatives for future studies.

Ultimately, the policy aim to prioritise specific interventions based on cost-effectiveness neglects the fact that strong therapeutic relationships with trusted clinicians are considerably more important for treatment success than the particular modality used. Thus, such comparisons are of limited value, whereas it could eventually be much more cost-effective to focus research and resources on improved training for clinicians instead.

### Lived Experience Commentary by Tamar Jeynes

It is interesting that in life much interacts with and mirrors itself. This systematic review endeavoured to be robust: 18 economic evaluations in 19.5 years fit the inclusion criteria. Nine different interventions, each with their own access criteria for service users.

Collecting robust economic data involves resource. This luxury is afforded to better funded interventions, which then make a case for future funding. Many service users do not meet criteria for these interventions.

It is interesting to identify what is missing.

There are no economic evaluations of survivor led or co-produced interventions, such as co-facilitation of therapies, survivor-led networks or crisis houses. These are more likely to not have inclusion criteria, reaching people that other interventions cannot. They often do not have the resource to conduct economic evaluations. Many have ceased to exist. Excluded service users only have access to costly emergency services during crisis. Being excluded depletes the internal resources needed to value their worth. Many cease to exist.

It is interesting that in life much interacts with and mirrors itself. When an intervention cannot demonstrate its worth, it cannot access funding which includes resource to measure its worth. When the only interventions that can demonstrate their worth are ones with inclusion criteria, excluded service users remain without a service. The cycle continues.

## Conclusions

There is no robust economic evidence to support a single intervention or model of community-based care for people with complex emotional needs that meet criteria for ‘personality disorder’ diagnoses. In line with clinical evidence, the review identified the strongest economic evidence for Dialectical Behavioural Therapy with all three identified studies indicating the intervention is likely to be cost-effective in community settings compared to treatment as usual. Consideration should be given to the limited availability of economic evaluations when interpreting this finding.

The studies included in this review were heterogeneous in terms of methods, outcome measurement (with 33 outcome measures used in the 18 studies), and the interventions studied. Future studies should aim to improve consistency in this field, and, given the paucity of evidence generated from small clinical trials, seek to evaluate existing and new services to provide ‘real world’ evidence on the cost and effects of therapies and services. Finally, whilst empirical evidence remains limited, economic modelling can support decisions on the best course of action.

## Data Availability

Data availability is not applicable to this article as no new data were created or analysed in this study.

## Contributions and Support

All authors contributed to the development of the methods and protocol. Searches, screening, data extraction, and quality appraisal were conducted by JB, TS, RS, AC and PM. JB, AC, SO, AS and PM drafted the manuscript. BLE, RS, SJ, EB, TJ and MC critically reviewed the manuscript. EB and TJ drafted the lived experience commentary. PM is the guarantor.

## Funding

This paper presents independent research commissioned and funded by the National Institute for Health Research (NIHR) Policy Research Programme, conducted by the NIHR Policy Research Unit (PRU) in Mental Health (Grant number: PR-PRU-0916-22003). The views expressed are those of the authors and not necessarily those of the NIHR, the Department of Health and Social Care or its arm’s length bodies, or other government departments.

EB and TJ are also authors of the independently written Lived experience commentary which follows the Discussion.

## Data Availability and Declarations

None of the authors have any competing interests to declare.

## Appendix Search Terms

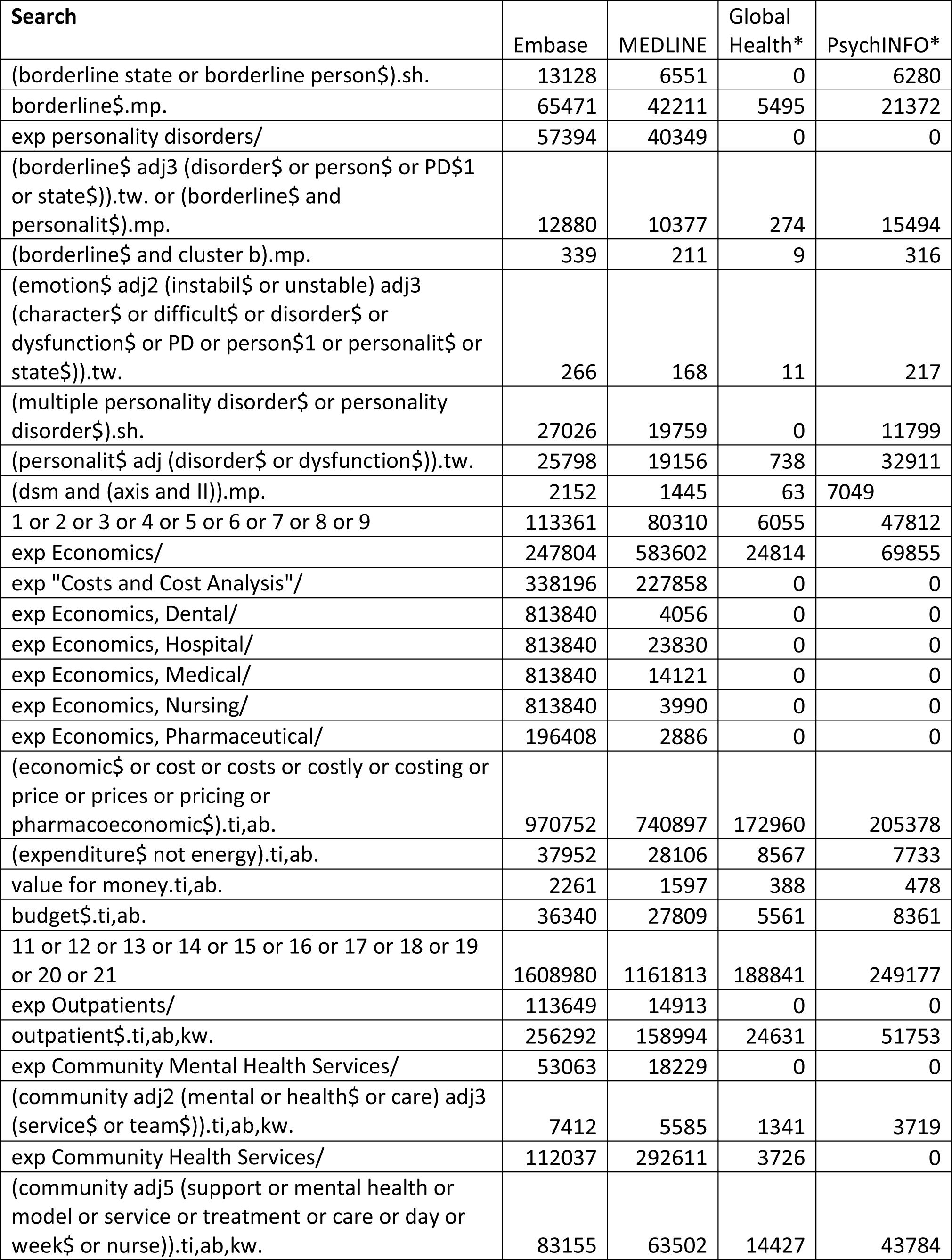
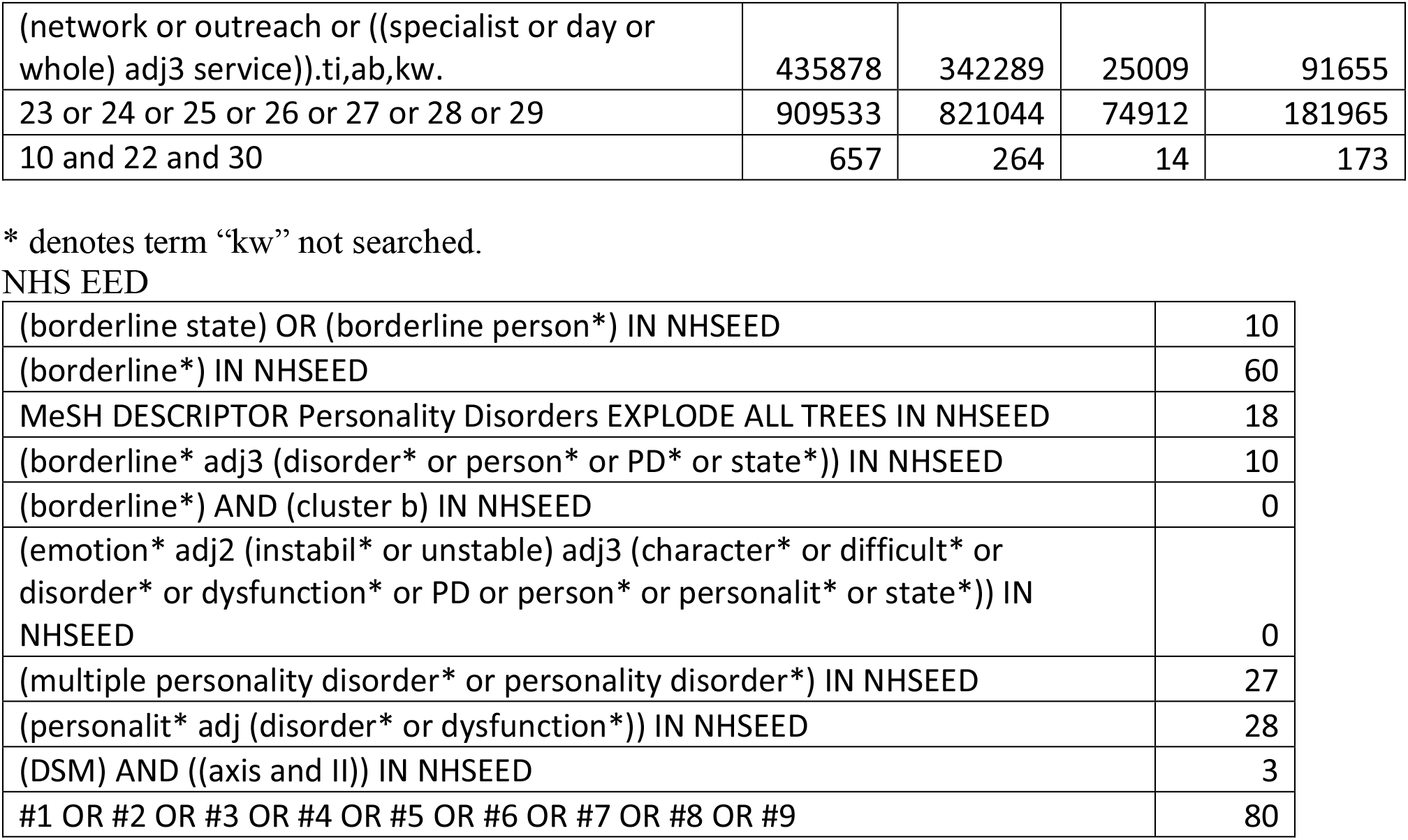

Search Terms
| Search | Embase | MEDLINE | Global Health* | PsychINFO* |
| --- | --- | --- | --- | --- |
| (borderline state or borderline person\$).sh. | 13128 | 6551 | 0 | 6280 |
| borderline\$.mp. | 65471 | 42211 | 5495 | 21372 |
| exp personality disorders/ | 57394 | 40349 | 0 | 0 |
| (borderline\$ adj3 (disorder\$ or person\$ or PD\$1 or state\$)).tw. or (borderline\$ and personalit\$).mp. | 12880 | 10377 | 274 | 15494 |
| (borderline\$ and cluster b).mp. | 339 | 211 | 9 | 316 |
| (emotion\$ adj2 (instabil\$ or unstable) adj3 (character\$ or difficult\$ or disorder\$ or dysfunction\$ or PD or person\$1 or personalit\$ or state\$)).tw. | 266 | 168 | 11 | 217 |
| (multiple personality disorder\$ or personality disorder\$).sh. | 27026 | 19759 | 0 | 11799 |
| (personalit\$ adj (disorder\$ or dysfunction\$)).tw. | 25798 | 19156 | 738 | 32911 |
| (dsm and (axis and II)).mp. | 2152 | 1445 | 63 | 7049 |
| 1 or 2 or 3 or 4 or 5 or 6 or 7 or 8 or 9 | 113361 | 80310 | 6055 | 47812 |
| exp Economics/ | 247804 | 583602 | 24814 | 69855 |
| exp "Costs and Cost Analysis"/ | 338196 | 227858 | 0 | 0 |
| exp Economics, Dental/ | 813840 | 4056 | 0 | 0 |
| exp Economics, Hospital/ | 813840 | 23830 | 0 | 0 |
| exp Economics, Medical/ | 813840 | 14121 | 0 | 0 |
| exp Economics, Nursing/ | 813840 | 3990 | 0 | 0 |
| exp Economics, Pharmaceutical/ | 196408 | 2886 | 0 | 0 |
| (economic\$ or cost or costs or costly or costing or price or prices or pricing or pharmacoeconomic\$).ti,ab. | 970752 | 740897 | 172960 | 205378 |
| (expenditure\$ not energy).ti,ab. | 37952 | 28106 | 8567 | 7733 |
| value for money.ti,ab. | 2261 | 1597 | 388 | 478 |
| budget\$.ti,ab. | 36340 | 27809 | 5561 | 8361 |
| 11 or 12 or 13 or 14 or 15 or 16 or 17 or 18 or 19 or 20 or 21 | 1608980 | 1161813 | 188841 | 249177 |
| exp Outpatients/ | 113649 | 14913 | 0 | 0 |
| outpatient\$.ti,ab,kw. | 256292 | 158994 | 24631 | 51753 |
| exp Community Mental Health Services/ | 53063 | 18229 | 0 | 0 |
| (community adj2 (mental or health\$ or care) adj3 (service\$ or team\$)).ti,ab,kw. | 7412 | 5585 | 1341 | 3719 |
| exp Community Health Services/ | 112037 | 292611 | 3726 | 0 |
| (community adj5 (support or mental health or model or service or treatment or care or day or week\$ or nurse)).ti,ab,kw. | 83155 | 63502 | 14427 | 43784 |
| (network or outreach or ((specialist or day or whole) adj3 service)).ti,ab,kw. | 435878 | 342289 | 25009 | 91655 |
| 23 or 24 or 25 or 26 or 27 or 28 or 29 | 909533 | 821044 | 74912 | 181965 |
| 10 and 22 and 30 | 657 | 264 | 14 | 173 |

\* denotes term "kw" not searched.

NHS EED
|  |  |
| --- | --- |
| (borderline state) OR (borderline person*) IN NHSEED | 10 |
| (borderline*) IN NHSEED | 60 |
| MeSH DESCRIPTOR Personality Disorders EXPLODE ALL TREES IN NHSEED | 18 |
| (borderline* adj3 (disorder* or person* or PD* or state*)) IN NHSEED | 10 |
| (borderline*) AND (cluster b) IN NHSEED | 0 |
| (emotion* adj2 (instabil* or unstable) adj3 (character* or difficult* or disorder* or dysfunction* or PD or person* or personalit* or state*)) IN NHSEED | 0 |
| (multiple personality disorder* or personality disorder*) IN NHSEED | 27 |
| (personalit* adj (disorder* or dysfunction*)) IN NHSEED | 28 |
| (DSM) AND ((axis and II)) IN NHSEED | 3 |
| #1 OR #2 OR #3 OR #4 OR #5 OR #6 OR #7 OR #8 OR #9 | 80 |

## References

1. Winsper C, Bilgin A, Thompson A, Marwaha S, Chanen AM, Singh SP, et al. The prevalence of personality disorders in the community: A global systematic review and meta-analysis. Vol. 216, British Journal of Psychiatry. Cambridge University Press; 2020. p. 69–78.

2. Shah R, Zanarini MC. Comorbidity of Borderline Personality Disorder: Current Status and Future Directions. Vol. 41, Psychiatric Clinics of North America. W.B. Saunders; 2018. p. 583–93.

3. Meuldijk D, McCarthy A, Bourke ME, Grenyer BFS. The value of psychological treatment for borderline personality disorder: Systematic review and cost offset analysis of economic evaluations. PLoS One. 2017 Mar 1;12(3).

4. Brettschneider C, Riedel-Heller S, König H-H. A Systematic Review of Economic Evaluations of Treatments for Borderline Personality Disorder. Yukich J, editor. PLoS One. 2014 Sep 29;9(9):e107748.

5. Beckwith H, Moran PF, Reilly J. Personality disorder prevalence in psychiatric outpatients: a systematic literature review. Vol. 8, Personality and mental health. Personal Ment Health; 2014. p. 91–101.

6. Storebø OJ, Stoffers-Winterling JM, Völlm BA, Kongerslev MT, Mattivi JT, Jørgensen MS, et al. Psychological therapies for people with borderline personality disorder. Vol. 2020, Cochrane Database of Systematic Reviews. John Wiley and Sons Ltd; 2020.

7. Oud M, Arntz A, Hermens MLM, Verhoef R, Kendall T. Specialized psychotherapies for adults with borderline personality disorder: A systematic review and meta-analysis. Vol. 52, Australian and New Zealand Journal of Psychiatry. SAGE Publications Inc., 2018. p. 949–61.

8. National Institute for Health and Care Excellence. Guide to the methods of technology appraisal 2013: Process and methods. 2013.

9. Drummond M, Sculpher M, Claxton K, Stoddart G, Torrance G. Methods for the Economic Evaluation of Health Care Programmes. 4th ed. Book. Oxford: Oxford University Press;

10. Shiell A, Donaldson C, Mitton C, Currie G. Health economic evaluation. Vol. 56, Journal of Epidemiology and Community Health. J Epidemiol Community Health; 2002. p. 85–8.

11. McCabe C, Claxton K, Culyer AJ. The NICE cost-effectiveness threshold: What it is and what that means. Vol. 26, PharmacoEconomics. Pharmacoeconomics; 2008. p. 733–44.

12. Moher D, Shamseer L, Clarke M, Ghersi D, Liberati A, Petticrew M, et al. Preferred reporting items for systematic review and meta-analysis protocols (PRISMA-P) 2015 statement. Syst Rev. 2015 Dec 1;4(1):1.

13. Husereau D, Drummond M, Petrou S, Carswell C, Moher D, Greenberg D, et al. Consolidated Health Economic Evaluation Reporting Standards (CHEERS) statement. BMJ. 2013 Mar 25;346:f1049.

14. Bamelis LLM, Arntz A, Wetzelaer P, Verdoorn R, Evers SMAA. Economic evaluation of schema therapy and clarification-oriented psychotherapy for personality disorders: A multicenter, randomized controlled trial. J Clin Psychiatry. 2015 Nov 1;76(11):e1432–40.

15. Grenyer BFS, Lewis KL, Fanaian M, Kotze B. Treatment of personality disorder using a whole of service stepped care approach: A cluster randomized controlled trial. PLoS One. 2018 Nov 1;13(11).

16. McMurran M, Crawford MJ, Reilly J, Delport J, McCrone P, Whitham D, et al. Psychoeducation with problem-solving (PEPS) therapy for adults with personality disorder: A pragmatic randomised controlled trial to determine the clinical effectiveness and cost-effectiveness of a manualised intervention to improve social functioning. Health Technol Assess (Rockv). 2016 Jul 1;20(52):1–249.

17. Kvarstein EH, Arnevik E, Halsteinli V, Rø FG, Karterud S, Wilberg T. Health service costs and clinical gains of psychotherapy for personality disorders: A randomized controlled trial of day-hospital-based step-down treatment versus outpatient treatment at a specialist practice. BMC Psychiatry. 2013 Nov 22;13.

18. Priebe S, Bhatti N, Barnicot K, Bremner S, Gaglia A, Katsakou C, et al. Effectiveness and Cost-Effectiveness of Dialectical Behaviour Therapy for Self-Harming Patients with Personality Disorder: A Pragmatic Randomised Controlled Trial. 2012;

19. Tyrer P, Milošeska K, Whittington C, Ranger M, Khaleel I, Crawford M, et al. Nidotherapy in the treatment of substance misuse, psychosis and personality disorder: Secondary analysis of a controlled trial. Psychiatrist. 2011 Jan;35(1):9–14.

20. Soeteman DI, Verheul R, Meerman AMMA, Ziegler U, Rossum B V., Delimon J, et al. Cost-effectiveness of psychotherapy for cluster C personality disorders: A decision-analytic model in The Netherlands. J Clin Psychiatry. 2011 Jan;72(1):51–9.

21. Soeteman DI, Verheul R, Delimon J, Meerman AMMA, Van Den Eijnden E, Rossum B V., et al. Cost-effectiveness of psychotherapy for cluster B personality disorders. Br J Psychiatry. 2010 May;196(5):396–403.

22. Blankers M, Koppers D, Laurenssen EMP, Peen J, Smits ML, Luyten P, et al. Mentalization-Based Treatment Versus Specialist Treatment as Usual for Borderline Personality Disorder: Economic Evaluation Alongside a Randomized Controlled Trial With 36-Month Follow-Up. J Pers Disord. 2019 Nov 4;1–20.

23. Borschmann R, Barrett B, Hellier JM, Byford S, Henderson C, Rose D, et al. Joint crisis plans for people with borderline personality disorder: Feasibility and outcomes in a randomised controlled trial. Br J Psychiatry. 2013 May;202(5):357–64.

24. Davidson KM, Tyrer P, Norrie J, Palmer SJ, Tyrer H. Cognitive therapy v. usual treatment for borderline personality disorder: Prospective 6-year follow-up. Br J Psychiatry. 2010 Dec;197(6):456–62.

25. Palmer S, Davidson K, Tyrer P, Gumley A, Tata P, Norrie J, et al. The cost-effectiveness of cognitive behavior therapy for borderline personality disorder: Results from the BOSCOT trial. J Pers Disord. 2006 Oct;20(5):466–81.

26. Pasieczny N, Connor J. The effectiveness of dialectical behaviour therapy in routine public mental health settings: An Australian controlled trial. Behav Res Ther. 2011 Jan;49(1):4–10.

27. Sinnaeve R, van den Bosch LMC, Hakkaart-van Roijen L, Vansteelandt K. Effectiveness of step-down versus outpatient dialectical behaviour therapy for patients with severe levels of borderline personality disorder: A pragmatic randomized controlled trial. Borderline Personal Disord Emot Dysregulation. 2018 Jul 10;5(1).

28. Van Asselt ADI, Dirksen CD, Arntz A, Giesen-Bloo JH, Van Dyck R, Spinhoven P, et al. Out-patient psychotherapy for borderline personality disorder: Cost-effectiveness of schema-focused therapy v. transference-focused psychotherapy. Br J Psychiatry. 2008 Jun;192(6):450–7.

29. Murphy A, Bourke J, Flynn D, Kells M, Joyce M. A cost-effectiveness analysis of dialectical behaviour therapy for treating individuals with borderline personality disorder in the community. Ir J Med Sci. 2019;

30. Tyrer P, Tom B, Byford S, Schmidt U, Jones V, Davidson K, et al. Differential effects of manual assisted cognitive behavior therapy in the treatment of recurrent deliberate self-harm and personality disturbance: The popmact study. Vol. 18, Journal of Personality Disorders. 2004. p. 102–16.

31. Ranger M, Tyrer P, Miloseska K, Fourie H, Khaleel I, North B, et al. Cost-effectiveness of nidotherapy for comorbid personality disorder and severe mental illness: Randomized controlled trial. Epidemiol Psichiatr Soc. 2009;18(2):128–36.

32. Tyrer P, Milošeska K, Whittington C, Ranger M, Khaleel I, Crawford M, et al. Nidotherapy in the treatment of substance misuse, psychosis and personality disorder: Secondary analysis of a controlled trial. Psychiatrist. 2011 Jan;35(1):9–14.

33. Bartak A, Spreeuwenberg MD, Andrea H, Holleman L, Rijnierse P, Rossum B V., et al. Effectiveness of different modalities of psychotherapeutic treatment for patients with cluster C personality disorders: Results of a large prospective multicentre study. Psychother Psychosom. 2009 Jan;79(1):20–30.

34. Bartak A, Andrea H, Spreeuwenberg MD, Ziegler UM, Dekker J, Rossum B V., et al. Effectiveness of outpatient, day hospital, and inpatient psychotherapeutic treatment for patients with cluster B personality disorders. Psychother Psychosom. 2011;80(1):28–38.

35. van Asselt ADI, Dirksen CD, Arntz A, Giesen-Bloo JH, Severens JL. The EQ-5D: A useful quality of life measure in borderline personality disorder? Eur Psychiatry. 2009 Mar;24(2):79–85.

36. Brazier J, Connell J, Papaioannou D, Mukuria C, Mulhern B, Peasgood T, et al. A systematic review, psychometric analysis and qualitative assessment of generic preference-based measures of health in mental health populations and the estimation of mapping functions from widely used specific measures. Health Technol Assess (Rockv). 2014;18(34):1–188.

37. ICHOM. Personality Disorders Standard Set [Internet]. [cited 2020 Sep 28]. Available from: https://www.ichom.org/portfolio/personality-disorders/

